# A systematic review and meta-analysis of Anakinra, Sarilumab, Siltuximab and Tocilizumab for Covid-19

**DOI:** 10.1101/2020.04.23.20076612

**Authors:** Fasihul Khan, Iain Stewart, Laura Fabbri, Samuel Moss, Karen A. Robinson, Alan Smyth, Gisli Jenkins

## Abstract

**Background:** There is accumulating evidence for an overly activated immune response in severe Covid-19, with several studies exploring the therapeutic role of immunomodulation. Through systematic review and meta-analysis, we assess the effectiveness of specific interleukin inhibitors for the treatment of Covid-19.

**Methods:** Electronic databases were searched on 7^th^ January 2021 to identify studies of immunomodulatory agents (anakinra, sarilumab, siltuximab and tocilizumab) for the treatment of Covid-19. The primary outcomes were severity on an ordinal scale measured at day 15 from intervention and days to hospital discharge. Key secondary endpoints included overall mortality.

**Results:** 71 studies totalling 22,058 patients were included, six were randomised trials. Most explored outcomes in patients who received tocilizumab (59/71). In prospective studies, tocilizumab was associated with improved unadjusted survival (RR 0.83 95%CI 0.72;0.96 I^2^ = 0.0%), but conclusive benefit was not demonstrated for other outcomes. In retrospective studies, tocilizumab was associated with less severe outcomes on an ordinal scale (Generalised odds ratio 1.34 95%CI 1.10;1.64, I^2^=98%) and adjusted mortality risk (HR 0.52 95%CI 0.41;0.66, I^2^ =76.6%). The mean difference in duration of hospitalisation was 0.36 days (95%CI -0.07;0.80, I^2^ =93.8%). There was substantial heterogeneity in retrospective studies, and estimates should be interpreted cautiously. Other immunomodulatory agents showed similar effects to tocilizumab, but insufficient data precluded meta-analysis by agent.

**Conclusion:** Tocilizumab was associated with a lower relative risk of mortality in prospective studies, but effects were inconclusive for other outcomes. Current evidence for the efficacy of anakinra, siltuximab or sarilumab in Covid-19 is insufficient, with further studies urgently needed for conclusive findings.

## INTRODUCTION

The novel severe acute respiratory syndrome coronavirus 2 (SARS-CoV-2) was first identified in Wuhan, China in December 2019(1). Since then, coronavirus disease 2019 (Covid-19) has been declared a global pandemic by the World Health Organisation (WHO) and continues to spread at an exponential rate with almost two million deaths reported worldwide (2, 3).

The clinical manifestations of Covid-19 tend to be heterogenous ranging from asymptomatic infection to acute respiratory disease syndrome (ARDS), multi-organ failure and death. Mechanisms underlying severe disease are incompletely understood, but accumulating evidence points towards a dysregulated and excessive host immune response referred to as cytokine storm syndrome (CSS)(4). During this state of immunological hyperactivation, increased circulating levels of pro-inflammatory cytokines including interleukin (IL)-1 and IL-6 have been demonstrated, and are associated with adverse clinical outcomes (5-7). Suppression of pro-inflammatory cytokines in Covid-19 may therefore be a potential therapeutic strategy (8).

SARS-CoV-2 shares a number of genetic and clinical similarities with other zoonotic coronaviruses, including severe acute respiratory syndrome coronavirus (SARS) and Middle East respiratory syndrome (MERS)(9, 10). There are also reports of elevated pro-inflammatory cytokines in patients with SARS and MERS (11, 12), suggesting overlapping therapeutic targets in the management of SARS, MERS and Covid-19.

Several clinical studies evaluating the role of immunomodulatory agents in Covid-19 have been published recently. Through systematic review and critical appraisal of the literature, we assess the effectiveness and safety of specific IL-1 (anakinra) and IL-6 (tocilizumab, siltuximab, sarilumab) inhibitors for the treatment of Covid-19, drawing on literature from previous similar coronavirus infections (SARS and MERS) where available. These agents already carry approval for the treatment of other rare non-infectious and autoimmune conditions, with an acceptable safety profile.

## METHODS

The systematic review was conducted in accordance with a pre-specified protocol (PROSPERO registration number: CRD42020176375), and has been reported in accordance with PRISMA (Preferred Reporting Items for Systematic Reviews and Meta-Analyses) guidelines(13).

### Search strategy and study selection

Electronic database searches were carried out in MEDLINE (1946 to latest) and EMBASE (1974 to latest), and ongoing clinical trial registries (clinicaltrials.gov and EU Clinical Trials Register), with the last search carried out on 7^th^ January 2021. Search terms were broad and included keywords and controlled vocabulary for patient and treatment-related terms (Figure S1 for MEDLINE search strategy). Unpublished and ongoing studies were identified by searching pre-print servers including medRxiv and bioRxiv. Searches were carried out independently by two reviewers in a standardised manner, followed by screening through titles and abstracts, before full text review. Disagreements were resolved by consensus, with unresolved conflicts decided by a third reviewer.

The review included all original studies excluding case reports, evaluating the use of at least one of anakinra, tocilizumab, sarilumab or siltuximab in patients aged over 18 with either suspected or confirmed Covid-19, SARS or MERS. Retrospective studies without a comparator arm were excluded due to their associated heterogeneity and inherent risk of bias. Language or year of publication restrictions were not applied. No minimal study sample size was specified for inclusion.

The planned primary outcomes were selected based on their clinical usefulness and included time to hospital discharge (days) and severity on an adapted four-point ordinal scale at day 15 following intervention, with the following ratings: i) death; ii) requirement for invasive mechanical ventilation (IMV) or Extra Corporal Membrane Oxygenation (ECMO); iii) hospitalised but no requirement for IMV/ECMO; iv) not hospitalised. Secondary outcomes included time to clinical improvement (days), duration of mechanical ventilation (days), overall mortality, mortality at 28 days and treatment related adverse events. For all outcomes studied, baseline was defined as the day of intervention.

### Data extraction and risk of bias assessment

Data were extracted from article text and figures using a data-extraction proforma and verified by a second reviewer. Information sought included study design, sample size, participant demographics, clinical investigation findings, intervention characteristics (name of agent, dose, route), treatment related adverse events, requirement and duration of invasive and non-invasive ventilation, use and dosage of oxygen, duration of hospital stay, survival outcome measures and follow up duration. Where ordinal outcomes were reported at multiple timepoints, those closest to day 15 post intervention were chosen for extraction. For ongoing trial protocols, the registration number, sample size, and expected date of completion were recorded.

Risk of bias assessment was carried out independently in duplicate. Due to the heterogeneity of study designs, various quality assessment tools available through the National Institute of Health were applied(14). The tools assess risk of bias through criterion specific to each study design, before providing an overall quality rating of good, fair or poor. Randomised studies were assessed using the Cochrane risk-of-bias tool for randomised trials (RoB2)(15). As per the review protocol, all studies were included irrespective of their risk of bias rating. Using the GRADE approach, we rated the overall quality of evidence for each outcome as high, moderate, low or very low(16).

### Statistical analysis

All identified studies were included in the narrative summary with summary tables for characteristics. For the primary outcomes, numbers of individuals meeting each outcome on the adapted ordinal scale were pooled using rank-based Wilcoxon Mann Whitney tests with ties split evenly between positive and negative outcomes, providing a generalised odds ratio (GenOR) with 95% confidence intervals (CI). The GenOR provides a measure of the likelihood that the intervention leads to a better rather than worse outcome when compared to a randomly chosen control (17). Mean hospital duration and standard deviation (SD) were extracted or were estimated from median and range/interquartile range (IQR) using the Box-Cox method (18). Mean difference in hospital stay was calculated where a control arm was reported. Where available, adjusted hazard ratios (HR) and unadjusted mortality data were extracted for quantitative synthesis. Where data were not reported in a tabular format, values were extracted from plotted data using a digital plot analyser(19).

Where sufficient studies were identified for a specific immunomodulator, findings were assessed using random effects meta-analysis and presented as forest plots. Meta-analyses were grouped by retrospective and prospective design and presented on the same plots with no overall estimate. The I^2^ statistic was used to evaluate statistical heterogeneity. Although sample sizes were limited, we used pseudo-R^2^ from meta-regression to explore variability in heterogeneity owing to study design (single-centre or multicentre), non-peer reviewed manuscripts, concomitant use of steroids, route of drug administration (intravenous or subcutaneous), and day outcome measured. Publication bias was assessed using funnel plot analysis and Egger’s test. Prospective studies without a control arm were excluded from meta-analysis and presented either in the narrative summary or in tables. All analyses were performed using Stata v.16 (StataCorp, College Station, TX, USA).

## RESULTS

Search of the electronic databases (MEDLINE and EMBASE) on 7^th^ January 2021 yielded a total of 2585 studies, with a further 576 studies identified through preprint servers. Following removal of duplicates, screening and full text review, 71 articles published worldwide were shortlisted for inclusion (anakinra, n=6; tocilizumab, n=58; anakinra and tocilizumab, n=1; sarilumab and tocilizumab, n=1; sarilumab, n=4; siltuximab, n=1) (Figure 1). Sixty-two studies were published in peer-reviewed journals, with the remaining nine identified through preprint servers. All studies were performed in patients with Covid-19, with no suitable studies identified for SARS or MERS. Twenty-nine studies were prospective in design, with seventeen studies including a control group for comparison, of which six were randomised studies. The remaining 42 studies were retrospective studies with control arms. Included studies provided a total of 22,058 patients, of which 7328 (33%) received one of the therapies under review alongside standard of care (SOC), and 14730 (67%) received SOC alone. Individual study characteristics for the published studies are presented in Tables 1 and 2 (and Tables S1-S2)

**Table 1.** Included studies with study characteristics and sample size for treatment (Tx) and control group (control) shown. * non peer-reviewed preprint study; #, study investigating both anakinra and tocilizumab; A, anakinra; Sa, sarilumab; Si, siltuximab; T, tocilizumab

| Author, year | Drug | N, Tx/<br>Control | Study<br>country | Centre | Study design | Author,<br>year | Drug | N, Tx/<br>Control | Study<br>country | Centre | Study design | Author,<br>year | Drug | N,Tx/<br>Control | Study<br>country | Centre | Study design |
| --- | --- | --- | --- | --- | --- | --- | --- | --- | --- | --- | --- | --- | --- | --- | --- | --- | --- |
| Balkhair,<br>2020(26) | A | 45/24 | Oman | single<br>centre | Prospective<br>with control | Roumier,<br>2020(32) | T | 49/47 | France | single<br>centre | Prospective<br>with control | Kimmig,<br>2020(33) | T | 54/57 | USA | single<br>centre | Retrospective |
| Huet,<br>2020(29) | A | 52/44 | France | single<br>centre | Prospective<br>with control | Salama,<br>2020(22) | T | 249/<br>128 | USA | multi-centre | Double blind<br>RCT | Klopfenste<br>in,<br>2020(34) | T | 20/25 | France | single<br>centre | Retrospective |
| Kooistra,<br>2020(35) | A | 21/39 | Netherla<br>nds | multi-centre | Prospective<br>with control | Salvarani,<br>2020(36) | T | 60/63 | Italy | multi-centre | Open label RCT | Lewis,<br>2020(37) | T | 497/497 | USA | multi-centre | Retrospective |
| *Kyriazopoul<br>ou, 2020(28) | A | 130/130 | Greece | multi-centre | Prospective | *Sanchez-Montalva,<br>2020(38) | T | 82/0 | Spain | single<br>centre | Prospective | Martinez-Sanz,<br>2020(39) | T | 260/969 | Spain | multi-centre | Retrospective |
| Cauchois,<br>2020(24) | A | 12/10 | France | multi-centre | Retrospective | Sciascia,<br>2020(40) | T | 63/0 | Italy | multi-centre | Prospective | Narain,<br>2020(27) | T | 73/3076 | USA | multi-centre | Retrospective |
| Cavalli,<br>2020(25) | A | 29/16 | Italy | single<br>centre | Retrospective | Stone,<br>2020(21) | T | 161/<br>82 | USA | multi-centre | Double blind<br>RCT | Nasa,<br>2020(41) | T | 22/63 | India | multi-centre | Retrospective |
| Narain,<br>2020(27) | A | 57/3076 | USA | multi-centre | Retrospective | Strohbehn,<br>2020(42) | T | 32/41 | USA | single<br>centre | Phase 2<br>open label | Patel,<br>2020(43) | T | 60/1505 | USA | single<br>centre | Retrospective |
| Benucci,<br>2020(44) | Sa | 8/0 | Italy | single<br>centre | Prospective | Toniati,<br>2020(45) | T | 100/0 | Italy | single<br>centre | Prospective | * Petrak,<br>2020 (46) | T | 81/37 | USA | multi-centre | Retrospective |
| Della-Torre,<br>2020(30) | Sa | 28/28 | Italy | single<br>centre | Prospective<br>with control | Biran,<br>2020(47) | T | 210/<br>420 | USA | multi-centre | Retrospective | Pettit,<br>2020(48) | T | 42/41 | USA | single<br>centre | Retrospective |
| * Gordon,<br>2021 (20) | Sa | 45/397 | UK | multi-centre | Adaptive RCT | Canziani,<br>2020(49) | T | 64/64 | Italy | multi-centre | Retrospective | Potere,<br>2020(50) | T | 74/74 | Italy | single<br>centre | Retrospective |
| Gremese,<br>2020(51) | Sa | 53/0 | Italy | single<br>centre | Prospective | Capra, 2020<br>(52) | T | 62/23 | Italy | single<br>centre | Retrospective | *Ramaswa<br>my,<br>2020(53) | T | 10/10 | USA | multi-centre | Retrospective |
| Sinha,<br>2020(54) | Sa | 255/0 | USA | single<br>centre | Prospective | Chillmuri,<br>2020(55) | T | 83/<br>685 | USA | single<br>centre | Retrospective | Rodriguez-Bano,<br>2020(56) | T | 21/65 | Spain | multi-centre | Retrospective |
| *Gritti<br>2020(31) | Si | 30/30 | Italy | single<br>centre | Prospective<br>with control | De Rossi,<br>2020(57) | T | 90/68 | Italy | single<br>centre | Retrospective | Rojas-Marté,<br>2020(58) | T | 88/344 | USA | single<br>centre | Retrospective |
| Albertini, 2020(59) | T | 22/22 | France | single centre | Prospective with control | Eimer, 2020(60) | T | 22/22 | Sweden | single centre | Retrospective | Roomi, 2020(61) | T | 96/97 | USA | single centre | Retrospective |
| Antony, 2020(62) | T | 80/0 | USA | multi-centre | Prospective | Fisher, 2020(63) | T | 45/70 | USA | single centre | Retrospective | Rosas, J.(64) | T | 20/17 | Spain | single centre | Retrospective |
| Campins, 2020(65) | T | 58/0 | Spain | single centre | Prospective | Galvan Roman, 2020(66) | T | 58/88 | Spain | single centre | Retrospective | Rossi, 2020(67) | T | 84/84 | France | single centre | Retrospective |
| *Carvalho, 2020(68) | T | 29/24 | Brazil | single centre | Prospective with control | *Garcia, 2020(69) | T | 77/94 | Spain | single centre | Retrospective | Rossotti, 2020(70) | T | 74/148 | Italy | single centre | Retrospective |
| Dastan, 2020(71) | T | 42/0 | Iran | single centre | Prospective | Gokhale, 2020(72) | T | 70/91 | India | single centre | Retrospective | Ruiz-Antoran, 2020(73) | T | 268/238 | Spain | multi-centre | Retrospective |
| *Gordon, 2021(20) | T | 350/397 | UK | multi-centre | Adaptive RCT | Guaraldi, 2020(74) | T | 179/365 | Italy | multi-centre | Retrospective | Somers, 2020(75) | T | 78/76 | USA | single centre | Retrospective |
| Hermine, 2020(23) | T | 63/67 | France | multi-centre | Open-label RCT | Guisado-Vasco, 2020(76) | T | 132/475 | Spain | single centre | Retrospective | Tian, 2020(77) | T | 65/130 | China | multi-centre | Retrospective |
| Malekzadeh, 2020(78) | T | 126/0 | Iran | multi-centre | Prospective | Gupta, 2020(79) | T | 433/3492 | USA | multi-centre | Retrospective | Tsai, 2020(80) | T | 66/66 | USA | single centre | Retrospective |
| Mikulska, 2020(81) | T | 29/66 | Italy | single centre | Prospective with control | Hill, 2020(82) | T | 43/45 | USA | single centre | Retrospective | *Wadud, 2020(83) | T | 84/84 | USA | single centre | Retrospective |
| Morena, 2020(84) | T | 51/0 | Italy | single centre | Prospective | Holt, 2020(85) | T | 24/30 | USA | single centre | Retrospective | Zheng, 2020(86) | T | 92/89 | China | single centre | Retrospective |
| Perrone 2020(87) | T | 708/481 | Italy | multi-centre | Single arm open label & Validation | Ip, 2020(88) | T | 134/413 | USA | multi-centre | Retrospective |  |  |  |  |  |  |
| *Rosas, 2020(89) | T | 294/144 | USA | multi-centre | Double blind RCT | Kewan, 2020(90) | T | 28/23 | USA | single centre | Retrospective |  |  |  |  |  |  |

**Table 2.** Treatment related adverse events. Adverse events for drug under study reported. Adverse events for control population reported where applicable. * non peer-reviewed preprint study

| Author, year | Therapy | Adverse effects |
| --- | --- | --- |
| Balkhair, 2020 | Anakinra | Treatment: infection (11%), ALT rise (14%). Control: infection (18%), ALT rise (9%) |
| Huet, 2020 | Anakinra | Treatment: ALT rise (13%). Control: 9% in anakinra |
| Kooistra, 2020 | Anakinra | Treatment: secondary infection (33%). Control: secondary infection (23%) |
| * Kyriazopoulou, 2020 | Anakinra | Increased leukopenia in treatment group vs. controls (8.5% vs 2.3%; p=0.05) |
| Cauchois, 2020 | Anakinra | N/R |
| Cavalli, 2020 | Anakinra | Treatment: staphylococcus epidermis (14%); deranged liver enzymes (10%). Control: bacteraemia (13%); deranged liver enzymes (31%). |
| Narain, 2020 | Anakinra | N/R |
| Benucci, 2020 | Sarilumab | Nil |
| Della-Torre, 2020 | Sarilumab | Treatment: Infections (21%); neutropenia (14%); liver enzyme increase (14%); thromboembolism (7%). Control: Infections (18%); thromboembolism (7%) |
| * Gordon, 2021 | Sarilumab | No serious event in sarilumab group, and 11 events in control |
| Gremese, 2020 | Sarilumab | neutropenia (15%); elevated liver enzymes (11%) |
| Sinha, 2020 | Sarilumab or Tocilizumab | bacterial infection (13%) |
| * Gritti 2020 | Siltuximab | Nil |
| Albertini, 2020 | Tocilizumab | elevated liver enzymes (64%) |
| Antony, 2020 | Tocilizumab | N/R |
| Campins, 2020 | Tocilizumab | Nil |
| * Carvalho, 2020 | Tocilizumab | Nil |
| Chillmuri, 2020 | Tocilizumab | N/R |
| Dastan, 2020 | Tocilizumab | transient diplopia (4.8%); Bell's palsy (2.4%) |
| * Gordon, 2021 | Tocilizumab | 9 serious adverse events in Tocilizumab group and 11 events in control |
| Hermine, 2020 | Tocilizumab | Treatment: Serious adverse events occurred in 20 (32%). Control: 29 (43%) (P=0.21) |
| Lewis, 2020 | Tocilizumab | Increased infection rate in treatment group (aOR 4.18; 95% CI 2.72-6.52) |
| Malekzadeh, 2020 | Tocilizumab | Nil |
| Mikulska, 2020 | Tocilizumab | N/R |
| Morena, 2020 | Tocilizumab | elevated liver enzymes (29%), thrombocytopenia (14%), neutropenia (6%), infections (24%) |
| Nasa, 2020 | Tocilizumab | two patients (9.1%) developed deranged LFTs and two patients (9.1%) developed secondary sepsis. |
| Perrone, 2020 | Tocilizumab | allergic reactions (0.4%), deranged liver enzymes (10.5%) |
| * Petrak, 2020 | Tocilizumab | N/R |
| * Rosas, I., 2020 | Tocilizumab | 66 serious infections (21%) were reported in the treatment arm and 49 (25.9%) in the placebo arm. Adverse events similar in both arms |
| Roumier, 2020 | Tocilizumab | Treatment: higher rates of neutropenia (35% vs. 0%, p<0.001). Control: trend towards increased bacterial infections (22% vs. 38%, p=0.089; including ventilator-acquired pneumonia: 8% vs. 26%, p=0.022) and shorter time to infection (mean 18 vs. 10 days, p=0.029) |
| Salama, 2020 | Tocilizumab | Serious adverse events occurred in 38 of 250 patients (15.2%) in the tocilizumab group and 25 of 127 patients (19.7%) in the placebo group. |
| Salvarani, 2020 | Tocilizumab | Nil |
| * Sanchez-Montalva, 2020 | Tocilizumab | Nil |
| Sciascia, 2020 | Tocilizumab | Nil |
| Stone, 2020 | Tocilizumab | Neutropenia developed in 22 patients in the treatment group, as compared with only 1 patient in the placebo group (P=0.002), but serious infections occurred in fewer patients in the tocilizumab group (13 [8.1%] vs. 14 [17.3%]; P=0.03). |
| Strohbehn, 2020 | Tocilizumab | Treatment: bacterial infections (15.6%). Control: not reported |
| Toniati, 2020 | Tocilizumab | septic shock (2%), gastrointestinal perforation (1%) |
| Biran, 2020 | Tocilizumab | Treatment: secondary bacterial infection in 17%. Control: secondary bacterial infection in 13% |
| Canziani, 2020 | Tocilizumab | HR 0.71 (95% CI 0.38-1.32) for infection; HR 0.89 (95% CI 0.39-2.06) for thrombosis; HR 1.17 (95% CI 0.47-2.92) for bleeding |
| Capra, 2020 | Tocilizumab | Nil |
| De Rossi, 2020 | Tocilizumab | Significant rise (from 44.3 +/- 28.3 to 103 +/- 141.3) in ALT in patients taking IV dose |
| Eimer, 2020 | Tocilizumab | Blood stream infection: 4 (18%) in treatment group vs. 6 (27%) in control |
| Fisher, 2020 | Tocilizumab | No increased risk of secondary infection (OR 1.17; 95%CI 0.51-2.71) |
| Galvan Roman, 2020 | Tocilizumab | N/R |
| * Garcia, 2020 | Tocilizumab | N/R |
| Gokhale, 2020 | Tocilizumab | N/R |
| Guaraldi, 2020 | Tocilizumab | 13% treated diagnosed with new infections vs 4% in control (p<0.0001) |
| Guisado-Vasco, 2020 | Tocilizumab | N/R |
| Gupta, 2020 | Tocilizumab | Treated and control patients experienced the following adverse events: secondary infection (140 [32.3%] vs 1085 [31.1%]); AST or ALT level elevation of more than 250 U/L (72 [16.6%] vs 452 [12.9%]) |
| Hill, 2020 | Tocilizumab | In treatment vs control group, there was increased sepsis (21% and 16%), ALT rise (9% vs 4%) and thrombocytopenia (12% vs 4%) |
| Holt, 2020 | Tocilizumab | N/R |
| Ip, 2020 | Tocilizumab | N/R |
| Kewan, 2020 | Tocilizumab | Similar rates of hospital-acquired infections occurred in both cohorts (18% in treatment and 22% in control). |
| Kimmig, 2020 | Tocilizumab | Treatment associated with increased secondary bacterial (aOR 2.76; 95% CI 1.11-7.2) and fungal (5.6% vs. 0%, p=0.112) infections |
| Klopfenstein, 2020 | Tocilizumab | N/R |
| Martinez-Sanz, 2020 | Tocilizumab | N/R |
| Narain, 2020 | Tocilizumab | N/R |
| Patel, 2020 | Tocilizumab | N/R |
| Pettit, 2020 | Tocilizumab | Overall infection rate was similar (16.2% treatment vs. 17.5% control), but late on-set infections occurred in more treated patients (23% vs 8%; p=0.013). In treated, 26% experienced an increase to >5 times upper limit normal of LFTs |
| Potere, 2020 | Tocilizumab | Nil |
| * Ramaswamy, 2020 | Tocilizumab | N/R |
| Rodriguez-Bano, 2020 | Tocilizumab | secondary bacterial infection similar in both groups (treated 12.5% vs. 10.3% control; p=0.57) |
| Rojas-Marte, 2020 | Tocilizumab | Bacteraemia was more common in the control group (24% vs. 13%, P=0.043), while fungemia was similar for both (3% vs. 4%, P=0.72) |
| Roomi, 2020 | Tocilizumab | N/R |
| Rosas, J. 2020 | Tocilizumab | Nil |
| Rossi, 2020 | Tocilizumab | N/R |
| Rossotti, 2020 | Tocilizumab | infectious complication in 32.4% |
| Ruiz-Antoran, 2020 | Tocilizumab | 32.6% in treated vs. 30.3% in control had increase in liver enzymes. Bacteraemia in 1 patient (0.4%) |
| Somers, 2020 | Tocilizumab | higher rate of superinfection in treated group (54% vs 26%; p<0.001) |
| Tian, 2020 | Tocilizumab | Deranged LFTs in 14% of tocilizumab and 14% of control group |
| Tsai, 2020 | Tocilizumab | N/R |
| * Wadud, 2020 | Tocilizumab | N/R |
| Zheng, 2020 | Tocilizumab | N/R |

**Figure 1.**
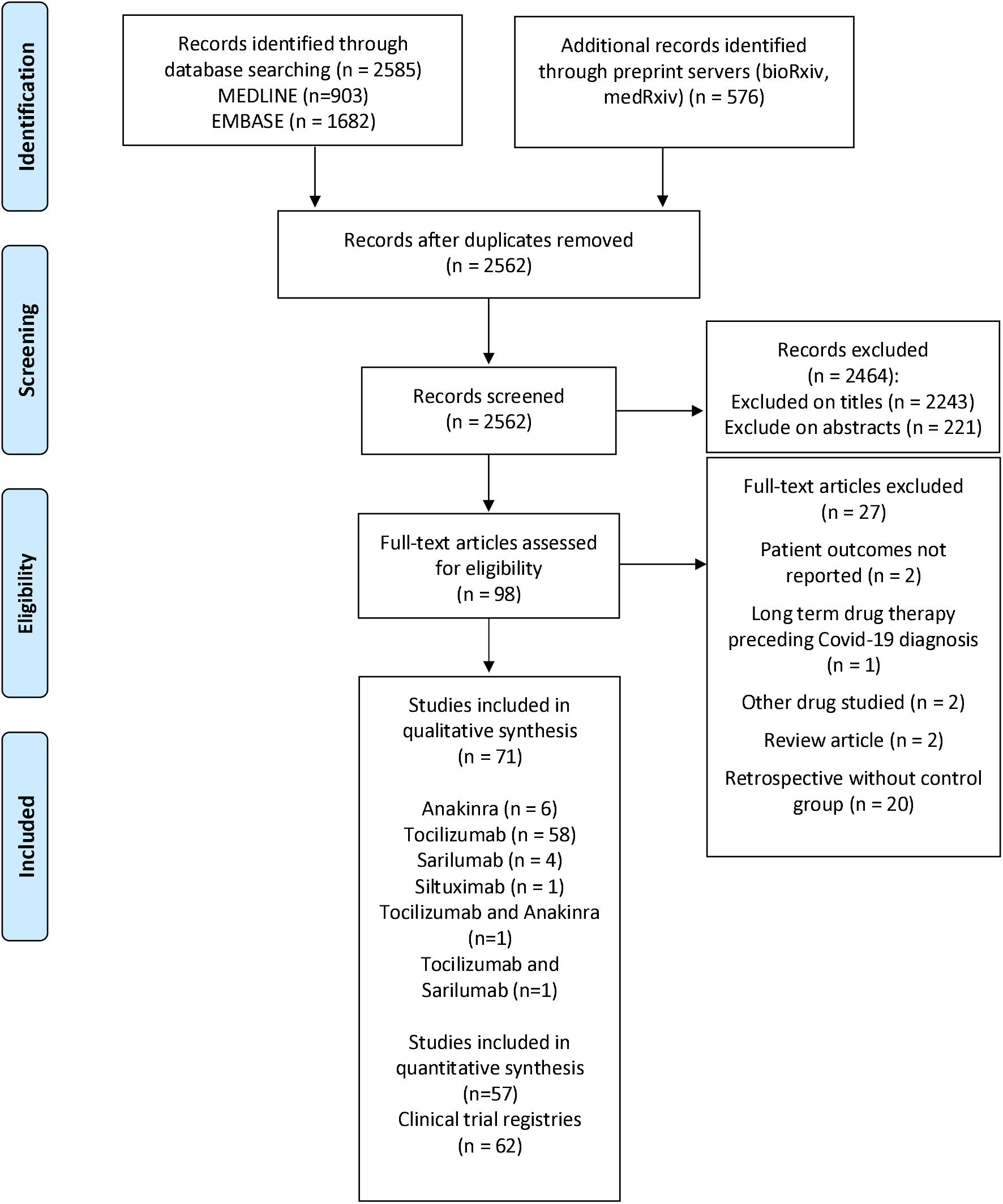
Flow diagram illustrates systematic search and screening strategy, including numbers meeting eligibility criteria and numbers excluded. Last search carried out on 7^th^ January 2021

Risk of bias assessment of the retrieved studies identified multiple limitations and highlighted a number of biases (Figure 2 and Table S3). The majority of included studies defined the study population specifically with clear inclusion/exclusion criteria. Where applicable, control participants were selected from the same population. However, many studies provided insufficient detail of the interventions and outcomes being studied or reporting was inconsistent, with key design, and outcome details omitted. Statistical analysis was variably reported, with few studies providing a sample size justification. In nearly all studies, patients were on concomitant therapies, limiting the ability to discern whether a specific intervention was related to the outcome. Following a formal risk of bias assessment, 23 (32%) studies were rated as good, 37 (52%) fair and 11 (15%) poor. Publication bias, assessed by observation of funnel plots and Egger’s test, was not present for any of the outcomes assessed (Figure S2).

**Figure 2.**
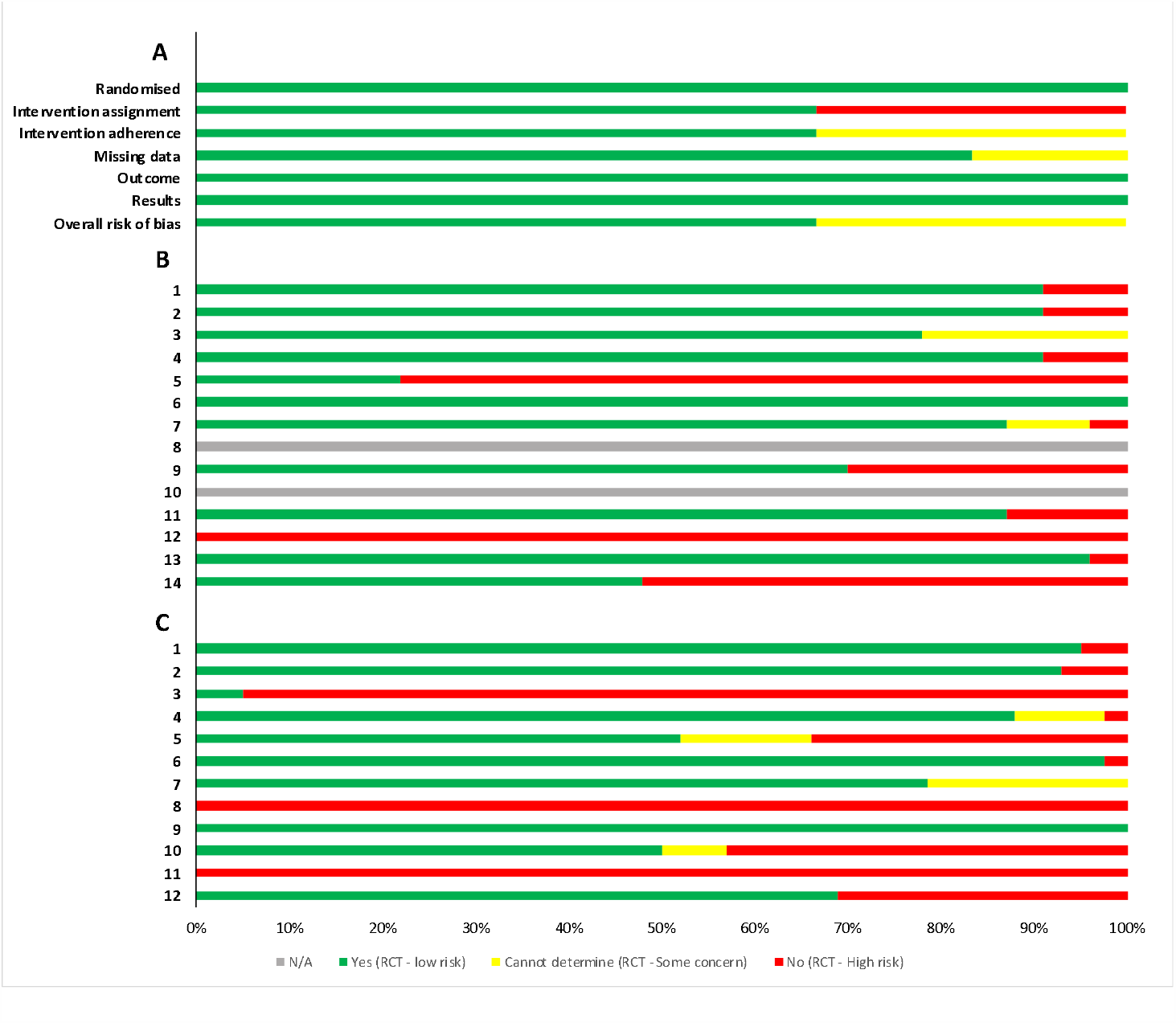
Summary of risk of bias assessment **A** - Randomised clinical trials assessed using Cochrane risk of bias 2 tool (n=6). Risk of bias was assessed in six categories and scored as either low risk of bias, some concern, or high risk of bias, before an overall risk of bias was given to each study. **B** - Non-randomised prospective studies (n=23). Questions numbered in the first column. 1. Was the research question or objective in this paper clearly stated? 2. Was the study population clearly specified and defined? 3. Was the participation rate of eligible persons at least 50%? 4. Were all the subjects selected or recruited from the same or similar populations (including the same time period)? Were inclusion and exclusion criteria for being in the study prespecified and applied uniformly to all participants? 5. Was a sample size justification, power description, or variance and effect estimates provided? 6. For the analyses in this paper, were the exposure(s) of interest measured prior to the outcome(s) being measured? 7. Was the timeframe sufficient so that one could reasonably expect to see an association between exposure and outcome if it existed? 8. For exposures that can vary in amount or level, did the study examine different levels of the exposure as related to the outcome (e.g., categories of exposure, or exposure measured as continuous variable)? 9. Were the exposure measures (independent variables) clearly defined, valid, reliable, and implemented consistently across all study participants? 10. Was the exposure(s) assessed more than once over time? 11. Were the outcome measures (dependent variables) clearly defined, valid, reliable, and implemented consistently across all study participants? 12. Were the outcome assessors blinded to the exposure status of participants? 13. Was loss to follow-up after baseline 20% or less? 14. Were key potential confounding variables measured and adjusted statistically for their impact on the relationship between exposure(s) and outcome(s)? **C** - Summary of risk of bias assessment for retrospective studies (n=42). Questions numbered in first column. 1. Was the research question or objective in this paper clearly stated and appropriate? 2. Was the study population clearly specified and defined? 3. Did the authors include a sample size justification? 4. Were controls selected or recruited from the same or similar population that gave rise to the cases (including the same timeframe)? 5. Were the definitions, inclusion and exclusion criteria, algorithms or processes used to identify or select cases and controls valid, reliable, and implemented consistently across all study participants? 6. Were the cases clearly defined and differentiated from controls? 7. If less than 100 percent of eligible cases and/or controls were selected for the study, were the cases and/or controls randomly selected from those eligible? 8. Was there use of concurrent controls? 9. Were the investigators able to confirm that the exposure/risk occurred prior to the development of the condition or event that defined a participant as a case? 10. Were the measures of exposure/risk clearly defined, valid, reliable, and implemented consistently (including the same time period) across all study participants? 11. Were the assessors of exposure/risk blinded to the case or control status of participants? 12. Were key potential confounding variables measured and adjusted statistically in the analyses? If matching was used, did the investigators account for matching during study analysis?

### Tocilizumab

Twelve prospective studies with a control arm, eight prospective studies without a control arm, and 40 retrospective studies examining the clinical impact of tocilizumab in Covid-19 were identified. Amongst the prospective studies there were six randomised clinical trials (RCTs). In total, the studies reported outcomes from 20,972 patients, of whom 6563 (31%) were given tocilizumab. Criteria for eligible participants varied across the studies, with many specifying respiratory failure with laboratory evidence of hyperinflammation as a prerequisite. The dose of tocilizumab administration was not entirely consistent with intravenous 8mg/kg or 400mg the most commonly studied route and dose.

#### Ordinal scale

A total of 12 studies provided outcomes on an adapted 4-point scale for 1782 patients including cases and controls (Table S4). The median time for reporting outcomes after treatment was 14 days (IQR 14-28). The recently available REMAP-CAP adaptive RCT interim analysis reported a signal that tocilizumab was associated with clinical improvement at day 14 (aOR 1.83 95%CI 1.40;2.41)(20), whilst in a separate RCT, outcomes on an ordinal severity scale did not differ between the treatment groups (HR 1.06 95%CI 0.80;1.41)(21). Distinctions in statistical methodology and clinical endpoints precluded inclusion of these RCT in the specified meta-analysis. Tocilizumab was not associated with better outcomes on the ordinal scale in meta-analysis of the remaining prospective studies, including three RCTs (GenOR 1.09 95% CI 0.99;1.19, I^2^ = 84.3%) (Figure 3). Variability in reported concomitant steroid administration had a significant contribution upon the substantial heterogeneity observed (Table S5). Tocilizumab was associated with better outcomes in meta-analysis of retrospective studies, indicating a 34% greater chance of less-severe outcomes on the adapted ordinal scale when compared to control (GenOR 1.34 95% CI 1.10;1.64, I^2^ = 98%). However, these results should be interpreted with caution as there was severe heterogeneity which could not be explained by variability in the factors assessed.

**Figure 3.**
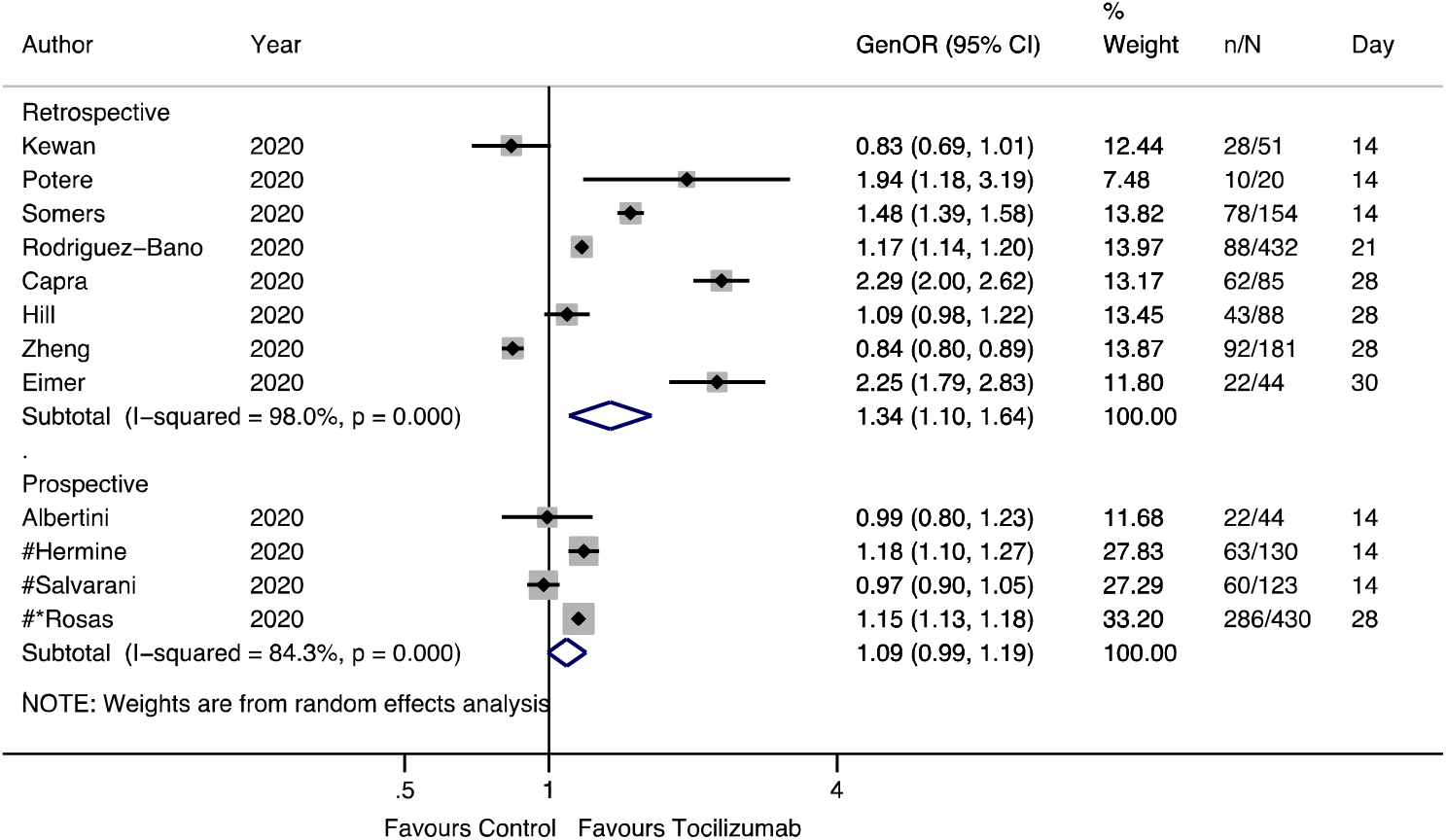
Tocilizumab generalised odds ratios (OR) for ordinal outcome forest plot. Generalised OR shown for each study with 95% confidence interval and day at which ordinal outcome recorded. Sample sizes given for patients receiving intervention (n) alongside total included (N) in study. Summary estimates presented separately for prospective and retrospective studies. * non peer-reviewed preprint studies # randomised controlled trials

#### Duration of hospitalisation

Two RCTs and nine retrospective studies reported the duration of hospitalisation for a total of 1553 survivors who received tocilizumab (Figure 4). Individual RCTs comparing the duration of hospitalisation with controls identified associations of tocilizumab with a reduced hospital stay (−0.34 days 95%CI -0.55;-0.12)(22) and earlier hospital discharge (aHR 1.41 95%CI 1.18;1.70)(20). Retrospective studies reporting the duration of hospitalisation were combined to give an overall summary estimate (20.98 days 95%CI 16.19;25.78, I^2^ = 97.1%), which was greater than the duration reported by RCTs (14.55 days 95%CI - 0.37;29.67, I^2^ = 99.9%). Compared with 943 patients in retrospective studies who received SOC only, tocilizumab was not associated with a difference in the mean duration of hospital stay (0.36 days 95% CI -0.07;0.80, I^2^ = 93.8%), with variability in route of administration (intravenous or subcutaneous) associated with the severe heterogeneity in this estimate (R^2^ = 81.64%, p<0.001).

**Figure 4.**
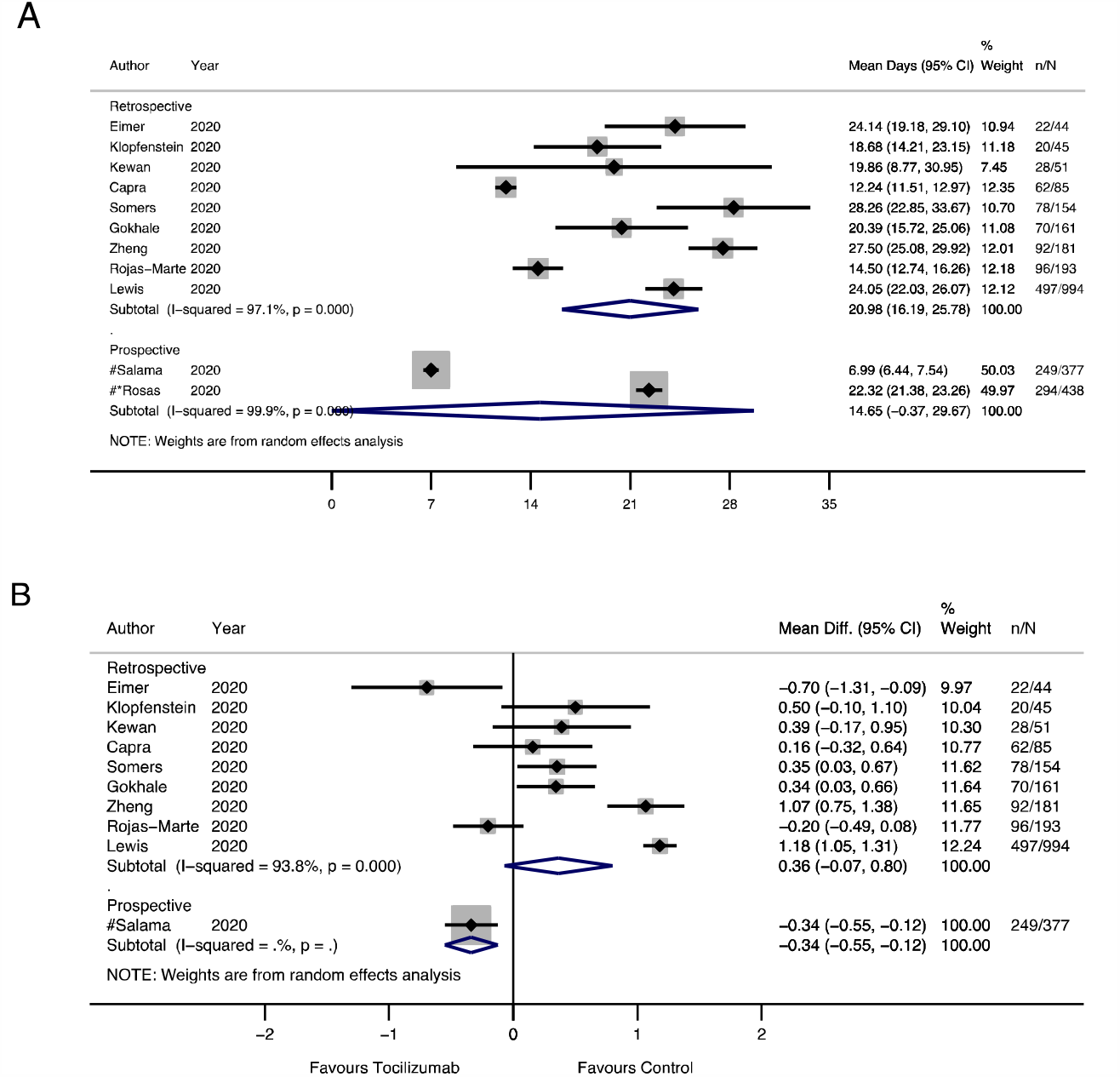
Tocilizumab duration of hospitalisation (days) forest plot. A: Mean duration of hospital stay. B: Mean difference compared with controls in duration of hospital stay. Effect sizes and associated 95% confidence intervals presented for each study. Sample sizes given for patients receiving intervention (n) and total included in study (N). Summary estimates presented separately for prospective and retrospective studies.

#### Overall mortality

Twenty-two studies totalling 13,702 patients reported adjusted hazard ratios for overall mortality, at a follow up time censored at a median of 28 days (IQR 14-30). Amongst the studies, two were RCTs and neither reported a difference between tocilizumab and control for mortality (21, 23). When prospective tocilizumab studies were pooled there was an emerging survival benefit, but the estimate was inconclusive (HR 0.70 95%CI 0.44;1.10, I^2^ = 0%) (Figure 5). In the remaining retrospective studies, tocilizumab was associated with a 48% lower risk of adjusted mortality with substantial heterogeneity (HR 0.52 95%CI 0.41;0.66, I^2^ = 76.6%). Meta-regression identified day outcome measured as a significant source of heterogeneity (R^2^ = 99.99, p=0.08).

**Figure 5.**
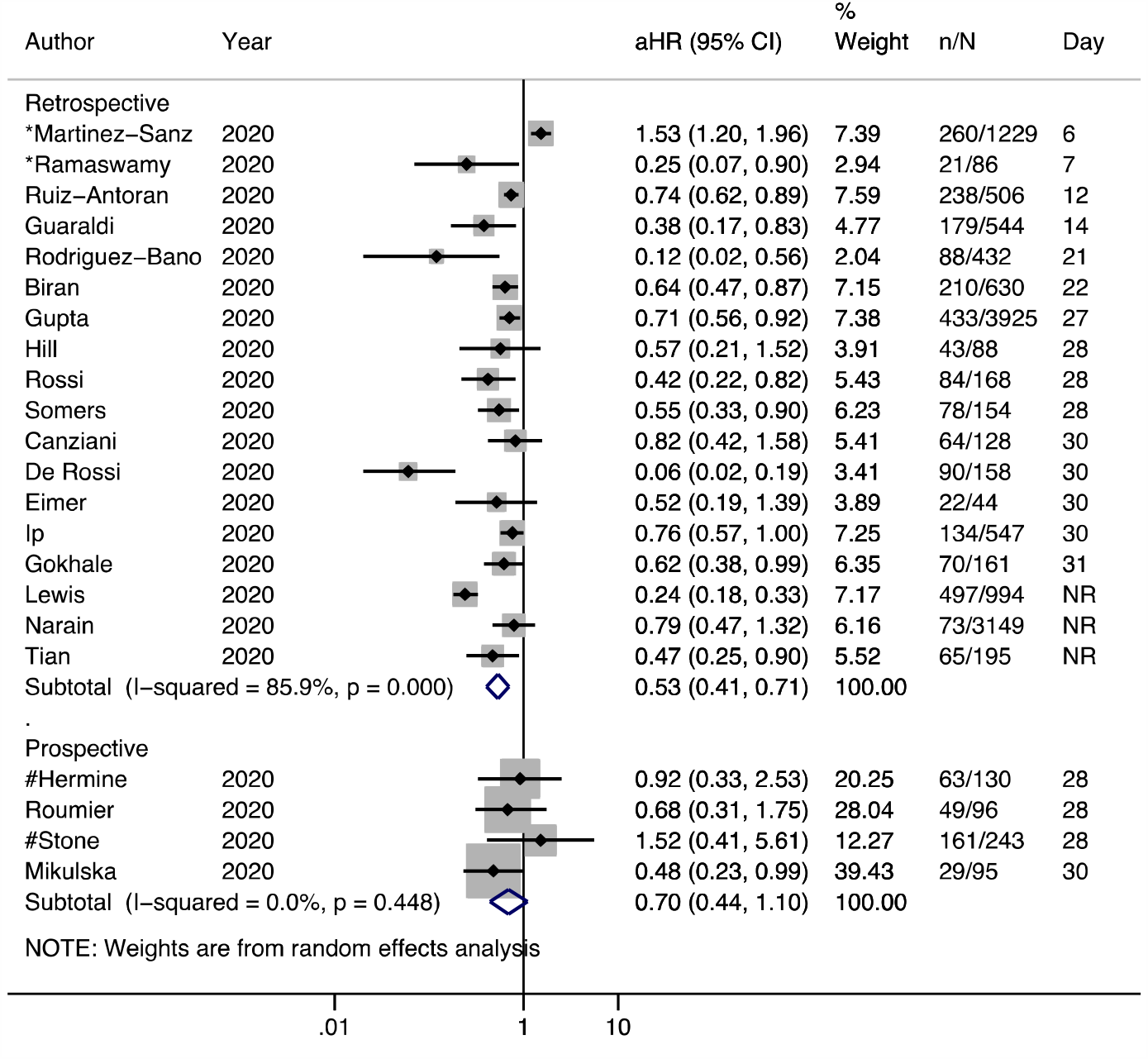
Tocilizumab adjusted hazard ratios (HR) for overall mortality forest plot. Adjusted HRs with associated 95% confidence interval and day of censorship presented for each study. Sample sizes given for patients receiving intervention (n) and total included (N) in study. Summary estimates presented separately for prospective and retrospective studies. * non peer-reviewed preprint studies # randomised controlled trials NR, not reported

Risk ratios (RR) were calculated from 42 studies, including six RCTs, reporting unadjusted mortality data for 15,085 patients at a median follow up of 24 days (IQR 14-28) (Figure 6). Tocilizumab was associated with a 17% lower unadjusted risk of mortality compared with the control arm in prospective studies (RR 0.83 95%CI 0.72;0.96, I^2^ = 0.0%), which did not reach significance in RCTs alone (RR 0.85 95%CI 0.71;1.01 I^2^ = 0.0%) (Figure S3). Within retrospective studies, tocilizumab was associated with a 24% lower risk of mortality (RR 0.76 95%CI 0.64;0.92, I^2^ = 80.3%), although there was substantial heterogeneity which could not be explained by variability in the factors assessed. The combined case fatality (CFR) across all studies included in the meta-analysis was 21.2% (1118/5284) in the intervention arm and 31.1% (3049/9801) in the control arm. The CFR from single arm prospective studies unable to be included in meta-analysis was 17.8% (113/634).

**Figure 6.**
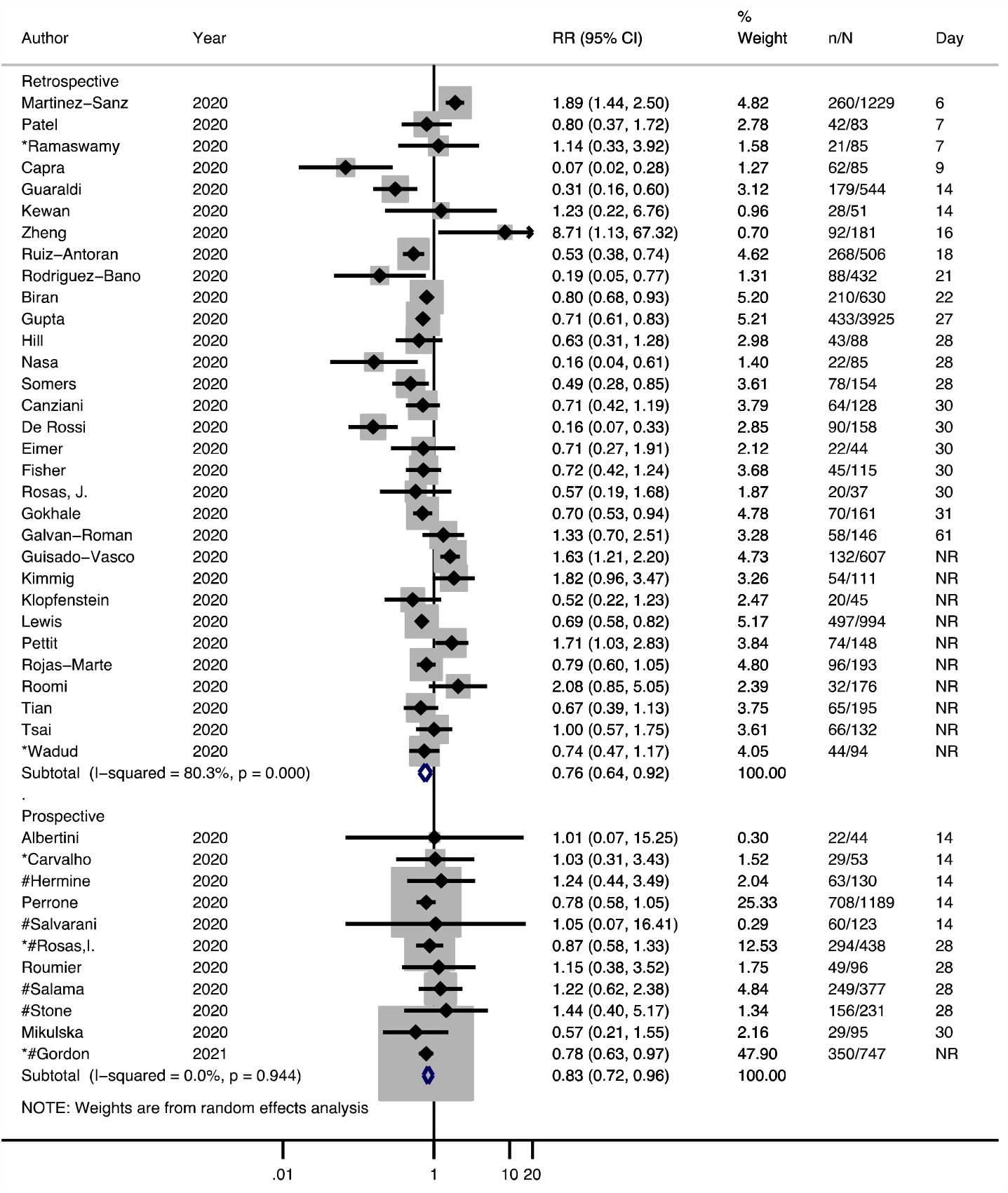
Tocilizumab mortality risk ratios (RR) forest plot. Risk ratios with associated 95% confidence interval and day of censorship presented for each study. Sample sizes given for patients receiving intervention (n) and total included in study (N). Summary estimates presented separately for prospective and retrospective studies. * non peer-reviewed preprint studies # randomised controlled trials NR, not reported

### Other immunomodulators

Studies exploring outcomes in patients who received anakinra, sarilumab or siltuximab were not quantitively synthesised for all outcomes, owing to differences in outcomes reported, study design and limited study numbers. Similar to studies in tocilizumab, participant criteria were inconsistent but typically included patients with respiratory failure and signs of hyperinflammation. Doses of therapeutic agents ranged from 200-600mg daily for anakinra, and 200-400mg daily for sarilumab. In all studies, patients received concomitant medications including but not limited to antivirals, hydroxychloroquine and corticosteroids. Meta-analysis inclusive of all immunomodulatory agents without sub analysis are presented in Figures S4-S7.

#### Anakinra

Four prospective and three retrospective studies exploring outcomes in 346 patients who received anakinra and 3339 controls were retrieved. Three studies reported ordinal outcome data for both anakinra and control participants, although the outcome day varied. Anakinra was associated with improved clinical outcomes in two retrospective studies of 22 and 45 patients, respectively (24, 25). A similar association with improved clinical outcomes was reported on day 14 in a prospective study of 69 patients (GenOR 1.77 95%CI 1.52;2.06)(26). Two studies reported adjusted HR for mortality with supportive results. A significant association was not observed in a retrospective study of 57 treated patients (aHR 0.79 95%CI 0.44;1.42)(27), whilst an association was observed in a prospective study of 130 patients (aHR 0.49 95%CI 0.26;0.91)(28). A significant unadjusted association was also observed in a further study of 52 patients treated with anakinra (HR 0.30; 95%CI 0.12-0.71)(29). Risk ratios were calculated from four studies totalling 424 participants. In a retrospective study of 29 treated patients, anakinra improved survival (RR 0.24 95%CI 0.07;0.79); associations were inconclusive when prospective studies were pooled (RR 0.70 95%CI 0.31;1.58, I^2^ = 32.8%) (Figure S8). No studies compared the duration of hospitalisation between recipients and non-recipients of anakinra.

#### Sarilumab

Five prospective studies exploring outcomes in 389 participants who received sarilumab were included. In the only RCT identified, sarilumab was associated with increased survival (aOR 2.01 95%CI 1.18;4.71), reduced duration of hospitalisation (aHR 1.60 95%CI 1.17;2.40) and improved ordinal outcomes at day 14 (aOR 1.86 95%CI 1.22;2.91)(20). In a further non-randomised study of 28 participants (30), sarilumab was not significantly associated with mortality (aHR 0.36 95%CI 0.08;1.68) and comparable effects were observed amongst treated and non-treated patients with respect to ordinal outcomes (GenOR 1.07 95%CI 0.90;1.27) and duration of hospitalisation (mean difference 0.02 95%CI -0.51;0.54). The combined CFR across the five included studies was 11% (43/389) for sarilumab, whilst in the only study reporting control mortality data the CFR was 35.8% (142/397).

#### Siltuximab

A single prospective cohort study of siltuximab studying outcomes in 60 patients was identified(31). Neither ordinal outcome data nor duration of hospitalisation were reported, but the adjusted risk of mortality was reported to be significantly lower in patients who received siltuximab (aHR 0.46 95%CI 0.22;0.97).

### Treatment related adverse events

Treatment related adverse events were reported in most studies (70%) and typically included secondary bacterial infections and derangement of liver enzymes (Table 2). In studies with a comparator arm exploring outcomes from patients who received anakinra or sarilumab, the frequency of treatment related adverse events was similar in both treatment and comparator groups. Findings from studies reporting outcomes following tocilizumab administration were inconsistent. In five studies, tocilizumab recipients had an increased prevalence of secondary infections compared with controls. However, in twelve studies, tocilizumab was associated with a lower or similar rate of secondary infections compared with controls.

### Clinical trials

Sixty-two planned or in-process clinical trials (tocilizumab, 44; siltuximab, 4; sarilumab, 9; anakinra, 13) were identified through clinical registry searches, with some clinical trials exploring more than one immunomodulatory agent. Currently registered clinical trials and their estimated dates of completion are provided in Figure S9.

## DISCUSSION

In this systematic review and meta-analysis, we summarise and evaluate the association between immunomodulatory agents and multiple outcomes in Covid-19. Although there was severe heterogeneity across tocilizumab studies exploring outcomes on an adapted four-point ordinal scale, a beneficial effect of tocilizumab was suggested in retrospective studies compared with controls. Prospective studies followed a similar direction of association, though confidence intervals were not conclusive. The certainty of the findings related to the adapted ordinal severity scale are assessed as moderate using GRADE (Table S6). The mean duration of hospitalisation was not altered by intervention, with low certainty of findings. Tocilizumab was associated with a survival benefit that was consistent across retrospective and prospective studies, with pooled analysis of unadjusted risk ratios demonstrating a 17% reduced risk of mortality in prospective studies. We assess the certainty of our findings related to overall mortality as high.

Due to heterogeneity in study designs and reported outcomes, studies in patients receiving immunomodulatory agents other than tocilizumab were not quantitatively synthesised for all outcomes. In the only study reporting adjusted HRs, anakinra was associated with reduced mortality. However, pooled analysis of unadjusted ratios in non-randomised studies did not demonstrate a mortality benefit. A single sarilumab RCT demonstrated that intervention was associated with improved outcomes and reduced hospital stay. No randomised studies were identified for siltuximab. For all agents included in this review, the frequency of adverse events was similar in the treatment and control arms. Sixty-one registered clinical trials exploring immunomodulatory agents in Covid-19 were identified, of which some have completed and been published.

In this review we highlight multiple limitations and considerable sources of inter-study heterogeneity. The majority of included studies were non-randomised cohorts of relatively modest size. Although most studies necessitated respiratory failure requiring at least basic respiratory support, participant criteria were not entirely consistent across the studies. The dosage and delivery of therapy varied across many of the non-randomised studies, and in nearly all studies patients were on concomitant medications such as antivirals, hydroxychloroquine and steroids with administration at the discretion of the treating physician, precluding causal associations of specific interleukin inhibitors with outcomes. Study outcomes were heterogeneous and a combination of clinical, laboratory and radiological outcomes were reported, rather than a single consistent endpoint. Furthermore, there was inconsistency in the duration of follow up and timing of reported outcomes. Individual patient data (IPD) may have mitigated some of these limitations, but in a rapidly progressing area, seeking IPD was deemed to be unrealistic due to the associated delays. We also observed significant statistical heterogeneity as measured by I^2^, and therefore the findings of our meta-analysis should be interpreted with caution. We were unable to explain all the residual heterogeneity using the factors we assessed, although concomitant steroid use, route of drug administration and day outcome measured appeared to contribute within specific outcomes.

To maximise value and timeliness of our review of four specific immunomodulators, two primary endpoints and a number of secondary endpoints, we included both retrospective and preprint studies. Risk of bias was minimised by restricting analysis of non-prospective studies to those with a control group, and caution is used to present summaries separately. We did not detect any significant publication bias in the reporting of effects. Where there was insufficient data for meta-analysis, summary outcomes were presented with qualitative synthesis to ensure the review was comprehensive. The data presented here represent findings from different countries, offering diversity in ethnic background. We were unable to identify suitable studies in SARS or MERS to comment on the generalisability of immunomodulators in other coronavirus outbreaks.

In conclusion, this systematic review provides the most up-to-date and complete evidence for a range of specific immunomodulatory therapies in the management of Covid-19. We have established that evidence for the efficacy of anakinra, siltuximab or sarilumab in Covid- 19 is currently insufficient and adequately powered high-quality randomised clinical studies are urgently needed. We demonstrate through quantitative synthesis of retrospective studies that tocilizumab intervention was frequently associated with improved outcomes and reduced mortality. However, data were highly heterogeneous and must be interpreted with caution. Prospective studies demonstrated a 17% lower unadjusted risk of mortality with tocilizumab, with minimal heterogeneity and similar adjusted estimates. Further research should focus on identifying participant and disease characteristics where immunomodulatory therapy is likely to be of maximal effectiveness, whilst also exploring the relationship with baseline inflammatory biomarkers such as interleukin-6 and C reactive protein. In summary, we demonstrate tocilizumab is associated with lower mortality in Covid-19, and other immunomodulatory therapies are worth exploring further.

## Data Availability

Not applicable

