## Supplementary for "A systematic review and meta-analysis of Anakinra, Sarilumab, Siltuximab and Tocilizumab for Covid-19"

#### Supplementary material

Figure 1 – MEDLINE search strategy

Figure 2 – Funnel plots for tocilizumab outcomes

Figure 3 – Tocilizumab forest plot for mortality risk ratios – RCTs only

Figure 4 – All agents forest plot for ordinal outcomes

Figure 5 – All agents forest plot for mean duration of hospitalisation

Figure 6 – All agents forest plot for mortality adjusted hazard ratios

Figure 7 – All agents forest plot for mortality risk ratios

Figure 8 - Anakinra mortality risk ratios (RR) forest plot

Figure 9 - Currently registered clinical trials

Table 1 – Characteristics of included studies

Table 2 – Patient characteristics and study outcomes

Table 3(a-c) – Risk of bias assessments

Table 4 – Primary outcome by individual study

Table 5 – Meta-regression values

Table 6 – GRADE rating

1. Respiratory Distress Syndrome, Adult/
2. SARS Virus/
3. Severe Acute Respiratory Syndrome/
4. severe acute respiratory distress syndrome*.mp.
5. Coronavirus Infections/
6. Coronavirus/
7. coronav*.mp.
8. covid*.mp.
9. SARS.mp.
10. Middle East Respiratory Syndrome Coronavirus/
11. MERS.mp.
12. anakinra.mp.
13. kineret.mp.
14. tocilizumab.mp.
15. altizumab.mp.
16. actemra.mp.
17. roactemra.mp.
18. sarilumab.mp.
19. kevzara.mp.
20. siltuximab.mp.
21. sylvant.mp.
22. Interleukin 1 Receptor Antagonist Protein/
23. anti-IL6.mp.
24. 1 or 2 or 3 or 4 or 5 or 6 or 7 or 8 or 9 or 10 or 11
25. 12 or 13 or 14 or 15 or 16 or 17 or 18 or 19 or 20 or 21 or 22 or 23
26. 24 and 25

**Supplementary Figure 1.** MEDLINE search strategy (last carried out on 7$^{th}$ January 2021)

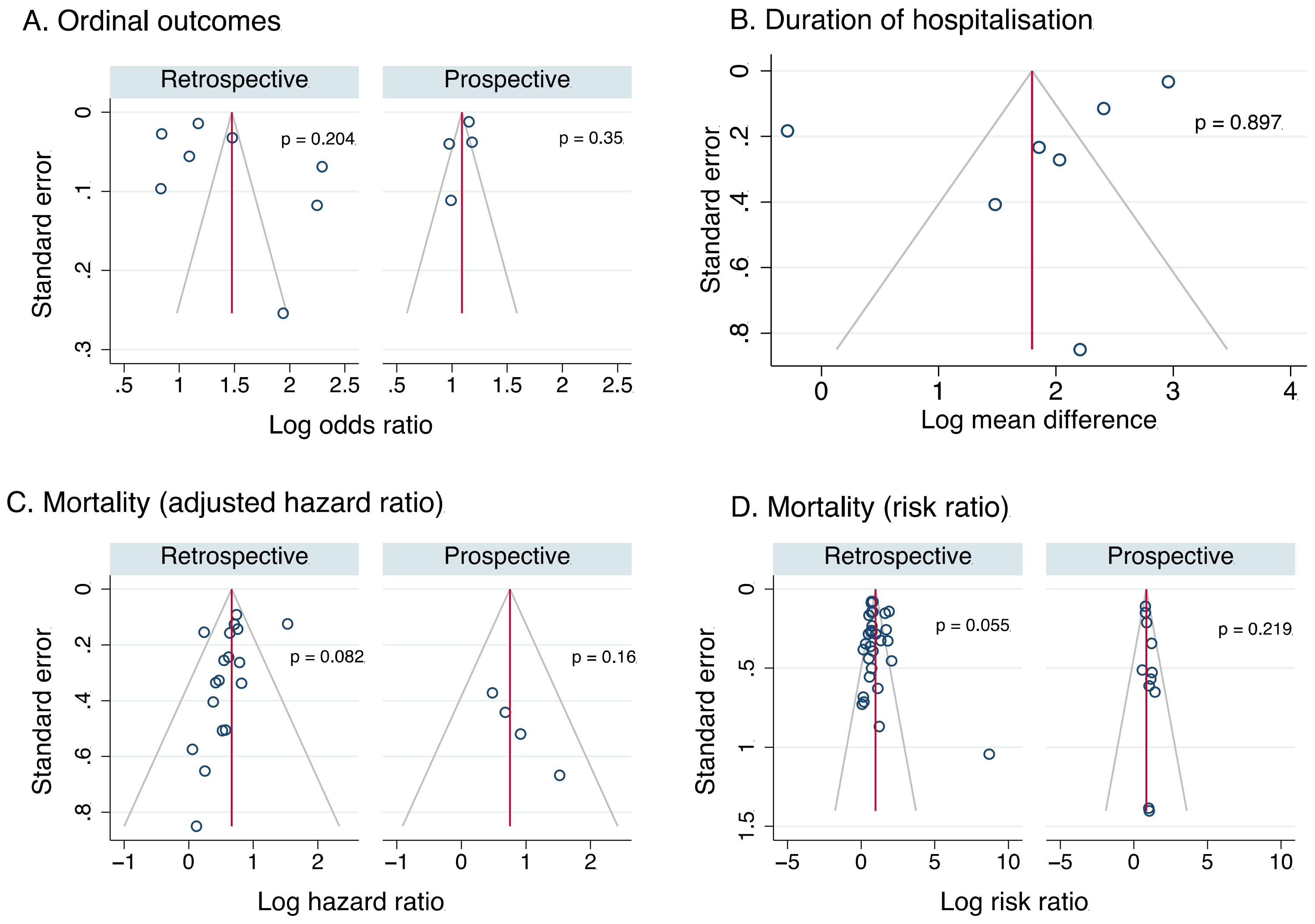

A. Ordinal outcomes
Retrospective
Prospective
Standard error
p = 0.204
p = 0.35
Log odds ratio
B. Duration of hospitalisation
p = 0.897
Log mean difference
C. Mortality (adjusted hazard ratio)
p = 0.082
p = 0.16
Log hazard ratio
D. Mortality (risk ratio)
p = 0.055
p = 0.219
Log risk ratio

**Supplementary Figure 2:** Funnel plots for outcomes evaluated in tocilizumab meta-analysis. A: ordinal outcomes, B: duration of hospitalisation, C: mortality (adjusted hazard ratio), D: mortality (risk ratio). Funnel plots presented separately for retrospective and prospective studies were applicable. Publication bias assessed using Egger's test, and p values presented next to funnel plot.

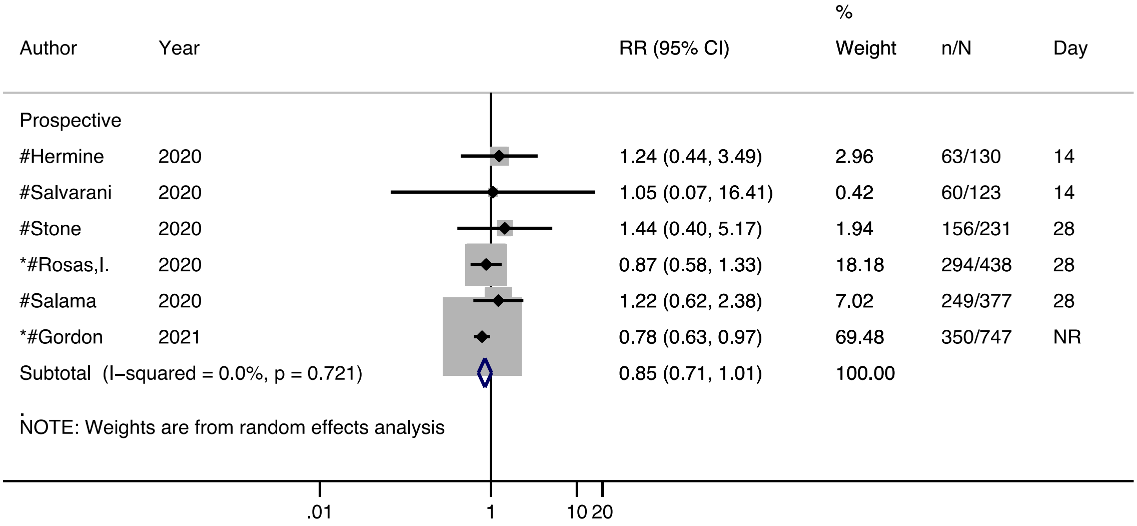

**Supplementary Figure 3** – Tocilizumab mortality risk ratios (RR) forest plot for randomised controlled trials only. Risk ratios with associated 95% confidence interval and day of censorship presented for each study. Sample sizes given for patients receiving intervention (n) and total included in study (N).
* non peer-reviewed preprint studies
### randomised controlled trials
NR, not reported

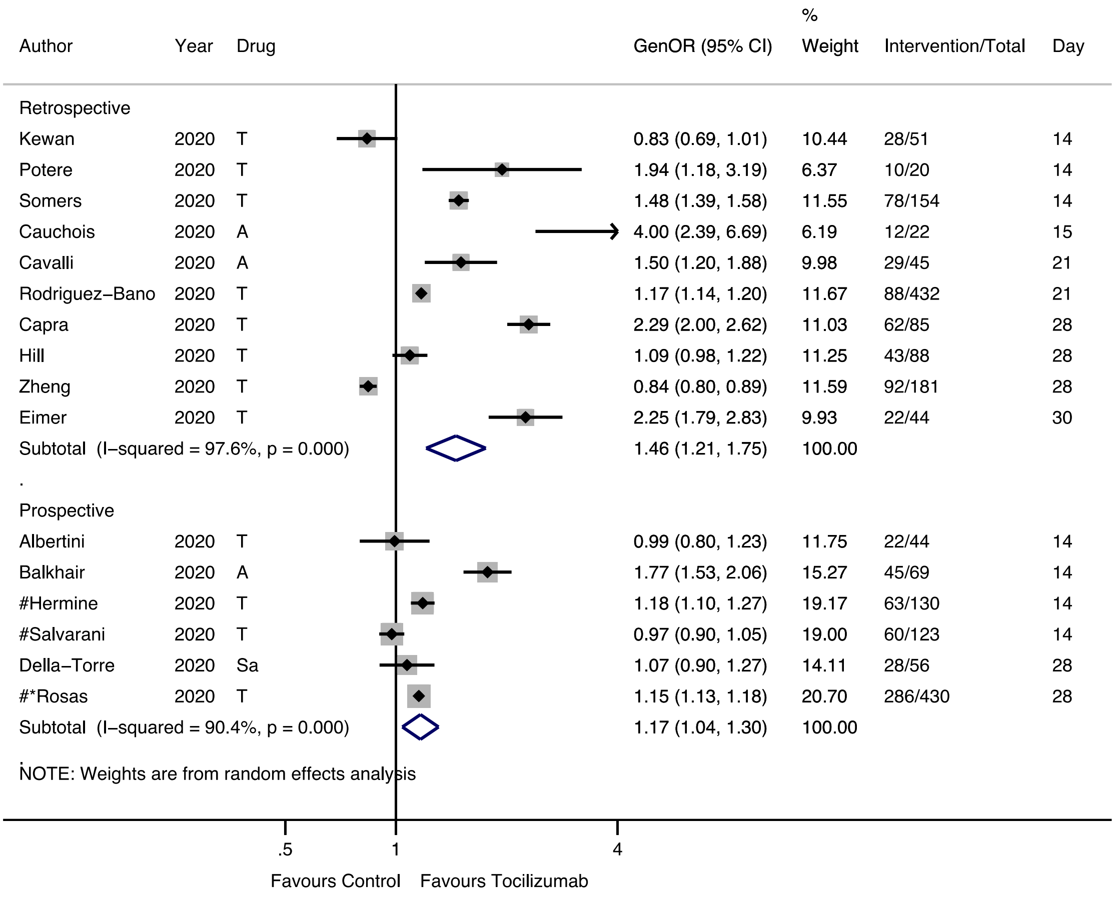

**Supplementary Figure 4** – All agents. Generalised odds ratios (OR) for ordinal outcome forest plot. Generalised OR shown for each study with 95% confidence interval and day at which ordinal outcome recorded. Sample sizes given for patients receiving intervention (n) alongside total included (N) in study. Summary estimates presented separately for prospective and retrospective studies. Drugs labelled where T = tocilizumab, A = anakinra, Sa = sarilumab

* non peer-reviewed preprint studies

### randomised controlled trials

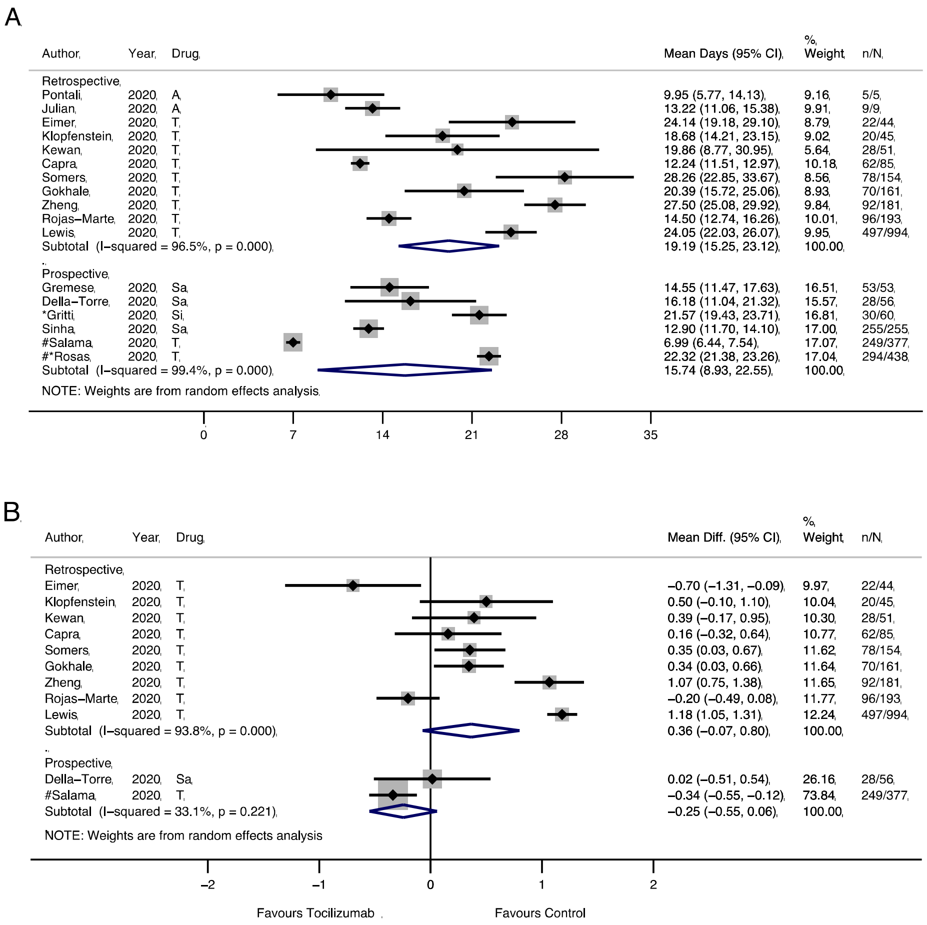

**Supplementary Figure 5** – All studies mean duration of hospitalisation (days) forest plot. **A**: Mean duration of hospital stay. **B**: Mean difference compared with controls in duration of hospital stay. Effect sizes and associated 95% confidence intervals presented for each study. Sample sizes given for patients receiving intervention (n) and total included in study (N). Summary estimates presented separately for prospective and retrospective studies. Drugs labelled where T = tocilizumab, Sa = sarilumab, Si = siltuximab.

* non peer-reviewed preprint studies

\# randomised controlled trials

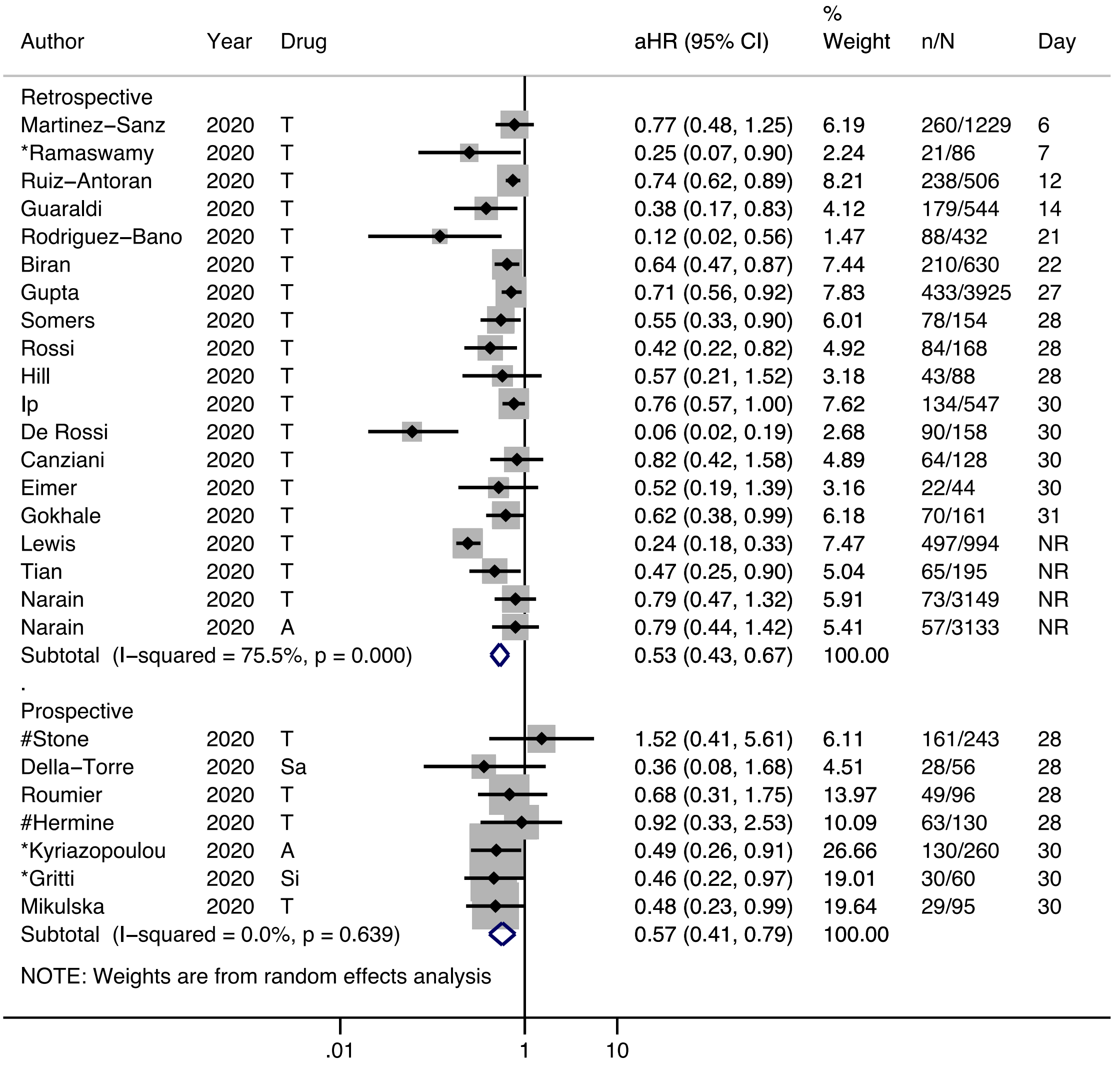

**Supplementary Figure 6** – All studies, adjusted hazard ratios (HR) for overall mortality forest plot. Adjusted HRs with associated 95% confidence interval and day of censorship presented for each study. Sample sizes given for patients receiving intervention (n) and total included (N) in study. Summary estimates presented separately for prospective and retrospective studies. Drugs labelled where T = tocilizumab, A = anakinra, Sa = sarilumab, Si = siltuximab.

* non peer-reviewed preprint studies

\# randomised controlled trials

NR, not reported

| Author | Year | Drug | RR (95% CI) | % Weight | n/N | Day |
|---|---|---|---|---|---|---|
| Retrospective | | | | | | |
| Martinez–Sanz | 2020 | T | 1.89 (1.44, 2.50) | 4.73 | 260/1229 | 6 |
| Patel | 2020 | T | 0.80 (0.37, 1.72) | 2.74 | 42/83 | 7 |
| *Ramaswamy | 2020 | T | 1.14 (0.33, 3.92) | 1.56 | 21/85 | 7 |
| Capra | 2020 | T | 0.07 (0.02, 0.28) | 1.26 | 62/85 | 9 |
| Guaraldi | 2020 | T | 0.31 (0.16, 0.60) | 3.08 | 179/544 | 14 |
| Kewan | 2020 | T | 1.23 (0.22, 6.76) | 0.95 | 28/51 | 14 |
| Zheng | 2020 | T | 8.71 (1.13, 67.32) | 0.70 | 92/181 | 16 |
| Ruiz–Antoran | 2020 | T | 0.53 (0.38, 0.74) | 4.54 | 268/506 | 18 |
| Cavalli | 2020 | A | 0.24 (0.07, 0.79) | 1.61 | 29/45 | 21 |
| Rodriguez–Bano | 2020 | T | 0.19 (0.05, 0.77) | 1.30 | 88/432 | 21 |
| Biran | 2020 | T | 0.80 (0.68, 0.93) | 5.09 | 210/630 | 22 |
| Gupta | 2020 | T | 0.71 (0.61, 0.83) | 5.10 | 433/3925 | 27 |
| Hill | 2020 | T | 0.63 (0.31, 1.28) | 2.94 | 43/88 | 28 |
| Nasa | 2020 | T | 0.16 (0.04, 0.61) | 1.39 | 22/85 | 28 |
| Somers | 2020 | T | 0.49 (0.28, 0.85) | 3.55 | 78/154 | 28 |
| Canziani | 2020 | T | 0.71 (0.42, 1.19) | 3.73 | 64/128 | 30 |
| De Rossi | 2020 | T | 0.16 (0.07, 0.33) | 2.81 | 90/158 | 30 |
| Eimer | 2020 | T | 0.71 (0.27, 1.91) | 2.10 | 22/44 | 30 |
| Fisher | 2020 | T | 0.72 (0.42, 1.24) | 3.63 | 45/115 | 30 |
| Rosas, J. | 2020 | T | 0.57 (0.19, 1.68) | 1.85 | 20/37 | 30 |
| Gokhale | 2020 | T | 0.70 (0.53, 0.94) | 4.69 | 70/161 | 31 |
| Galvan–Roman | 2020 | T | 1.33 (0.70, 2.51) | 3.24 | 58/146 | 61 |
| Guisado–Vasco | 2020 | T | 1.63 (1.21, 2.20) | 4.64 | 132/607 | NR |
| Kimmig | 2020 | T | 1.82 (0.96, 3.47) | 3.21 | 54/111 | NR |
| Klopfenstein | 2020 | T | 0.52 (0.22, 1.23) | 2.44 | 20/45 | NR |
| Lewis | 2020 | T | 0.69 (0.58, 0.82) | 5.06 | 497/994 | NR |
| Pettit | 2020 | T | 1.71 (1.03, 2.83) | 3.78 | 74/148 | NR |
| Rojas–Marte | 2020 | T | 0.79 (0.60, 1.05) | 4.70 | 96/193 | NR |
| Roomi | 2020 | T | 2.08 (0.85, 5.05) | 2.36 | 32/176 | NR |
| Tian | 2020 | T | 0.67 (0.39, 1.13) | 3.69 | 65/195 | NR |
| Tsai | 2020 | T | 1.00 (0.57, 1.75) | 3.56 | 66/132 | NR |
| *Wadud | 2020 | T | 0.74 (0.47, 1.17) | 3.98 | 44/94 | NR |
| Subtotal (I–squared = 80.2%, p = 0.000) | | | 0.75 (0.62, 0.90) | 100.00 | | |
| . | | | | | | |
| Prospective | | | | | | |
| Albertini | 2020 | T | 1.01 (0.07, 15.25) | 0.26 | 22/44 | 14 |
| Balkhair | 2020 | A | 1.33 (0.28, 6.37) | 0.79 | 45/69 | 14 |
| *Carvalho | 2020 | T | 1.03 (0.31, 3.43) | 1.34 | 29/53 | 14 |
| #Hermine | 2020 | T | 1.24 (0.44, 3.49) | 1.80 | 63/130 | 14 |
| *Kyriazopoulou | 2020 | A | 0.38 (0.15, 0.93) | 2.35 | 130/260 | 14 |
| Perrone | 2020 | T | 0.78 (0.58, 1.05) | 22.40 | 708/1189 | 14 |
| #Salvarani | 2020 | T | 1.05 (0.07, 16.41) | 0.26 | 60/123 | 14 |
| *Della–Torre | 2020 | Si | 0.40 (0.08, 1.89) | 0.80 | 28/56 | 28 |
| Kooistra | 2020 | A | 1.06 (0.35, 3.21) | 1.57 | 21/50 | 28 |
| *#Rosas,I. | 2020 | T | 0.87 (0.58, 1.33) | 11.08 | 294/438 | 28 |
| Roumier | 2020 | T | 1.15 (0.38, 3.52) | 1.55 | 49/96 | 28 |
| #Salama | 2020 | T | 1.22 (0.62, 2.38) | 4.28 | 249/377 | 28 |
| #Stone | 2020 | T | 1.44 (0.40, 5.17) | 1.18 | 156/231 | 28 |
| Mikulska | 2020 | T | 0.57 (0.21, 1.55) | 1.91 | 29/95 | 30 |
| *#Gordon | 2021 | T | 0.78 (0.63, 0.97) | 42.36 | 350/747 | NR |
| *#Gordon | 2021 | Sa | 0.84 (0.48, 1.48) | 6.06 | 45/442 | NR |
| Subtotal (I–squared = 0.0%, p = 0.908) | | | 0.82 (0.71, 0.94) | 100.00 | | |

NOTE: Weights are from random effects analysis

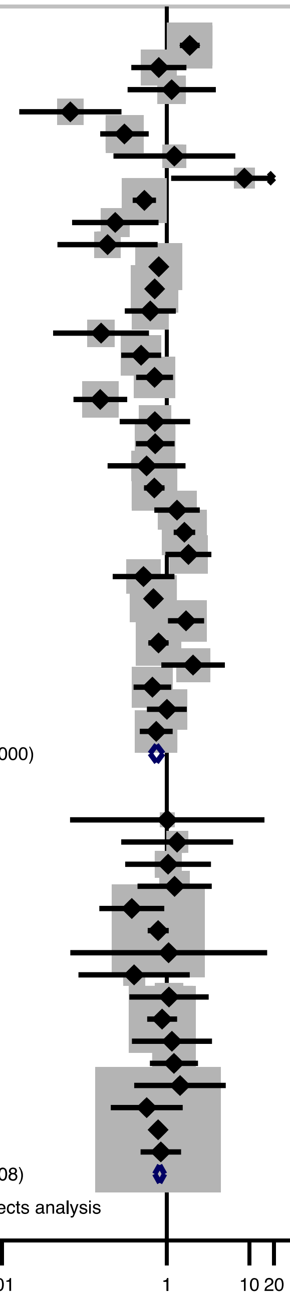

**Supplementary Figure 7** – All agents, mortality risk ratios (RR) forest plot. Risk ratios with associated 95% confidence interval and day of censorship presented for each study. Sample sizes given for patients receiving intervention (n) and total included in study (N). Summary estimates presented separately for prospective and retrospective studies. Drugs labelled where T = tocilizumab, A = anakinra, Si = siltuximab, Sa = sarilumab
* non peer-reviewed preprint studies
### randomised controlled trials
NR, not reported

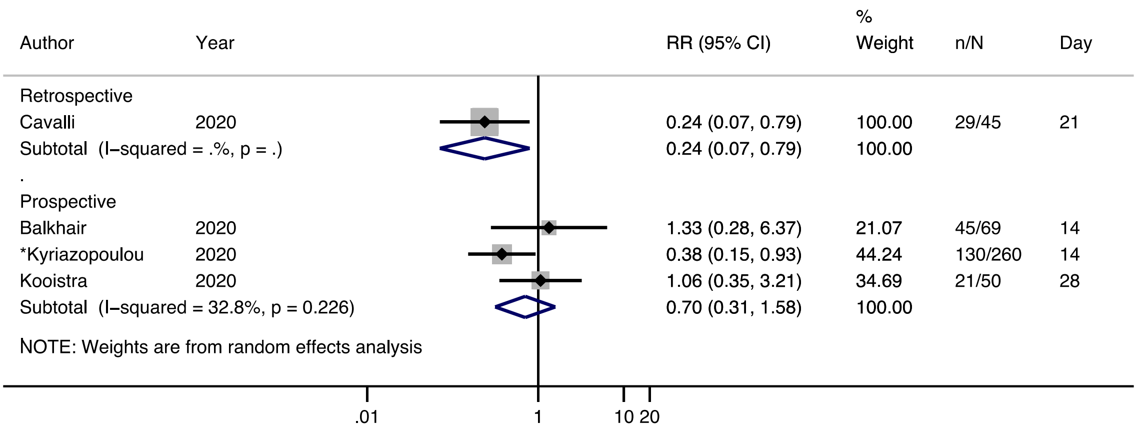

**Supplementary Figure 8** – Anakinra mortality risk ratios (RR) forest plot. Risk ratios with associated 95% confidence interval and day of censorship presented for each study. Sample sizes given for patients receiving intervention (n) and total included in study (N). Summary estimates presented separately for prospective and retrospective studies.

* non peer-reviewed preprint studies

**Estimated completion date (quarter)**

| Clinical Trial No. | Date | Sample size | 2020 | | | 2021 | | | | 2022 | | | |
|---|---|---|---|---|---|---|---|---|---|---|---|---|---|
| | | | Q2 | Q3 | Q4 | Q1 | Q2 | Q3 | Q4 | Q1 | Q2 | Q3 | Q4 |
| NCT04315480 | May-20 | 38 | | | | | | | | | | | |
| NCT04310228 | May-20 | 150 | | | | | | | | | | | |
| ChiCTR2000029765 | May-20 | 188 | | | | | | | | | | | |
| NCT04322188 | May-20 | 50 | | | | | | | | | | | |
| NCT04329650 | May-20 | 100 | | | | | | | | | | | |
| NCT04306705 | May-20 | 120 | | | | | | | | | | | |
| NCT04346355 | May-20 | 398 ‡ | | | | | | | | | | | |
| ChiCTR2000030196 | May-20 | 60 | | | | | | | | | | | |
| NCT04359667 | Jun-20 | 30 | | | | | | | | | | | |
| NCT04492501 | Jul-20 | 600 | | | | | | | | | | | |
| NCT04357860 | Jul-20 | 120 | | | | | | | | | | | |
| NCT04335305 | Aug-20 | 24 | | | | | | | | | | | |
| NCT04363736 | Aug-20 | 100 | | | | | | | | | | | |
| NCT04320615 | Aug-20 | 450 ‡ | | | | | | | | | | | |
| NCT04366232 | Aug-20 | 54 | | | | | | | | | | | |
| NCT04327388 | Aug-20 | 421 | | | | | | | | | | | |
| NCT04519385 | Aug-20 | 69 | | | | | | | | | | | |
| NCT04435717 | Aug-20 | 78 | | | | | | | | | | | |
| NCT04445272 | Aug-20 | 500 | | | | | | | | | | | |
| NCT04315298 | Aug-20 | 1912 | | | | | | | | | | | |
| NCT04462757 | Sep-20 | 5 | | | | | | | | | | | |
| NCT04364009 | Sep-20 | 240 | | | | | | | | | | | |
| NCT04372186 | Sep-20 | 379 ‡ | | | | | | | | | | | |
| NCT04335071 | Oct-20 | 100 | | | | | | | | | | | |
| NCT04345445 | Oct-20 | 310 | | | | | | | | | | | |
| NCT04356937 | Oct-20 | 300 ‡ | | | | | | | | | | | |
| NCT04332094 | Oct-20 | 276 | | | | | | | | | | | |
| NCT04361032 | Oct-20 | 260 | | | | | | | | | | | |
| NCT04560205 | Oct-20 | 50 | | | | | | | | | | | |
| NCT04377503 | Dec-20 | 40 | | | | | | | | | | | |
| NCT04409262 | Dec-20 | 450 | | | | | | | | | | | |
| NCT04386239 | Dec-20 | 40 | | | | | | | | | | | |
| NCT04330638 | Dec-20 | 342* | | | | | | | | | | | |
| NCT04324021 | Dec-20 | 54 | | | | | | | | | | | |
| NCT04357808 | Dec-20 | 30 | | | | | | | | | | | |
| NCT04341584 | Dec-20 | 240 | | | | | | | | | | | |
| NCT04412291 | Feb-21 | 120 * | | | | | | | | | | | |
| NCT04331795 | Mar-21 | 332 ‡ | | | | | | | | | | | |
| NCT04362111 | Mar-21 | 30 | | | | | | | | | | | |
| NCT04332913 | Mar-21 | 30 | | | | | | | | | | | |
| NCT04443881 | Mar-21 | 180 | | | | | | | | | | | |
| NCT04479358 | Mar-21 | 332 | | | | | | | | | | | |
| NCT04377750 | May-21 | 500 | | | | | | | | | | | |
| NCT04377659 | May-21 | 40 | | | | | | | | | | | |
| NCT04423042 | Jun-21 | 30 | | | | | | | | | | | |
| NCT04322773 | Jun-21 | 200* | | | | | | | | | | | |
| NCT04486521 | Jul-21 | 11000 * | | | | | | | | | | | |
| NCT04403685 | Jul-21 | 129 † | | | | | | | | | | | |
| NCT04363853 | Aug-21 | 200 | | | | | | | | | | | |
| NCT04364009 | Sep-21 | 240 | | | | | | | | | | | |
| NCT04324073 | Dec-21 | 239 | | | | | | | | | | | |
| NCT04331808 | Dec-21 | 228 ‡ | | | | | | | | | | | |
| NCT04476979 | Dec-21 | 120 | | | | | | | | | | | |
| NCT04412772 | Dec-21 | 300 | | | | | | | | | | | |
| NCT04339712 | Apr-22 | 40* | | | | | | | | | | | |
| NCT04359901 | Apr-22 | 120 | | | | | | | | | | | |
| NCT04357366 | Apr-22 | 100 | | | | | | | | | | | |
| NCT04370834 | Apr-22 | 217 † | | | | | | | | | | | |
| NCT04361552 | May-22 | 180 † | | | | | | | | | | | |
| NCT04424056 | Nov-22 | 216 * | | | | | | | | | | | |
| NCT04317092 | Dec-22 | 400 ‡ | | | | | | | | | | | |
| NCT02735707 | Dec-22 | 7100 * | | | | | | | | | | | |

| | |
|---|---|
| | Tocilizumab |
| | Siltuximab |
| | Sarilumab |
| | Anakinra |

**Supplementary Figure 9 -** Currently registered clinical trials with estimated completion date presented per calendar year quarter. Clinical trials are stratified as per colour key. * same study investigating multiple immunomodulatory agents. † study has been terminated. ‡ results available

(last search 5th Oct)

| Author, year | Study country | Centre | Study design | Dose | Participant criteria | Outcomes reported | Concomitant therapies |
|---|---|---|---|---|---|---|---|
| **ANAKINRA** | | | | | | | |
| Bakhair, 2020 | Oman | single centre | Prospective with control | 100mg S/C twice daily for 72h, then 100mg daily for 7 days | respiratory failure, bilateral lung infiltrates | mortality, ventilatory requirements | antibiotics |
| Huet, 2020 | France | single centre | Prospective with control | 100mg S/C twice daily for 72h, then 100mg daily for 7 days | respiratory failure, bilateral lung infiltrates | mortality, ventilatory requirement, laboratory biomarkers | hydroxychloroquine, antibiotics, IV methylprednisolone |
| Kooistra, 2020 | Netherlands | multi-centre | Prospective with control | 300mg IV then 100mg 6 hourly | IMV | mortality, ventilatory requirement, laboratory biomarkers | antivirals, hydroxychloroquine, corticosteroids |
| *Kyriazopoulou | Greece | multi-centre | Prospective | 100mg S/C daily for 10 days | lung infiltrates and suPAR level ≥6ug/L | respiratory failure, mortality, SOFA score | hydroxychloroquine, antivirals, antibiotics, corticosteroids |
| Cauchois, 2020 | France | multi-centre | Retrospective | 300mg IV daily for 5 days then tapered over 3 days | respiratory failure and CRP > 110mg/L | ventilatory requirement, laboratory biomarkers | hydroxychloroquine, antibiotics |
| Cavalli, 2020 | Italy | single centre | Retrospective | 10mg/kg/day IV | moderate-severe ARDS requiring CPAP and hyperinflammation | survival, ventilatory requirement, CRP | CPAP, hydroxychloroquine, lopinavir, ritonavir |
| Narain, 2020 | USA | multi-centre | Retrospective | N/R | hyperinflammation | hospital mortality | hydroxychloroquine |
| **SARILUMAB** | | | | | | | |
| Benucci, 2020 | Italy | single centre | Prospective | 400mg IV repeated twice at 200mg at 48 hourly intervals | N/R | ventilatory requirement, laboratory biomarkers | hydroxychloroquine, azithromycin, antivirals |
| Della-Torre, 2020 | Italy | single centre | Prospective with control | 400mg IV | radiological bilateral lung infiltrates and hyperinflammation | overall survival, ventilatory requirements | hydroxychloroquine, azithromycin, antivirals |
| * Gordon, 2021 | UK | multi-centre | Adaptive RCT | 400mg IV | within 24h of ICU admission with respiratory failure | respiratory and cardiovascular organ support-free days up to day 21, mortality, time to discharge | corticosteroids, remdesivir |

| | | | | | | | |
|---|---|---|---|---|---|---|---|
| Gremese, 2020 | Italy | single centre | Prospective | 400mg IV | respiratory failure and radiological infiltrates | ventilatory requirement, discharge from ICU, mortality | hydroxychloroquine, azithromycin, antivirals |
| Sinha, 2020 | USA | single centre | Prospective | 200mg IV | respiratory failure and hyperinflammation | mortality, discharge from hospital, IMV | hydroxychloroquine, azithromycin |
| **SILTUXIMAB** | | | | | | | |
| *Gritti 2020 | Italy | single centre | Prospective with control | 11mg/kg IV. Second dose 72 hours later (n=6) | respiratory failure requiring IVM or non-IVM support | mortality, time to IVM, laboratory biomarkers | antivirals, hydroxychloroquine, corticosteroids |
| **TOCILIZUMAB** | | | | | | | |
| Albertini, 2020 | France | single centre | Prospective with control | 8mg/kg IV. Second dose 72 hours later (n=20) | respiratory failure, bilateral radiological infiltrates, elevated CRP | respiratory rate, oxygen requirements, laboratory biomarkers | hydroxychloroquine and azithromycin |
| Antony, 2020 | USA | multi-centre | Prospective | 4mg/kg/day IV 12 hourly | supplemental oxygen dose >3L/min, but not mechanically ventilated | mortality, ventilatory requirement, laboratory biomarkers | methylprednisolone |
| Campins, 2020 | Spain | single centre | Prospective | N/R | N/R | mortality | corticosteroids (98%) |
| *Carvalho, 2020 | Brazil | single centre | Prospective with control | 400mg IV two doses | respiratory failure, hyperinflammation | in-hospital mortality, need for renal replacement therapy, inflammatory and oxygenation markers, use of antibiotics | hydroxychloroquine, azithromycin |
| Dastan, 2020 | Iran | single centre | Prospective | 400mg IV | severe: respiratory failure, or bilateral radiological infiltrates, IL-6>10pg/mL critical: need for ICU or IMV | oxygen requirements, ventilatory requirements, death, laboratory biomarkers | antivirals |
| * Gordon, 2021 | UK | multi-centre | Adaptive RCT | 8mg/kg IV repeated after 12-24h | within 24h of ICU admission with respiratory failure | respiratory and cardiovascular organ support-free days up to day 21, mortality, time to discharge | corticosteroids, remdesivir |

| | | | | | | | |
|---|---|---|---|---|---|---|---|
| Hermine, 2020 | France | multi-centre | Open label RCT | 8mg/kg IV | radiological infiltrates with respiratory failure but not admitted to ICU | dead or ventilatory support on day 4, survival at day 14, laboratory biomarkers | antivirals, corticosteroids |
| Malekzadeh, 2020 | Iran | multi-centre | Prospective | 324mg or 486mg SC (weight dependent) | respiratory failure and hyperinflammation | all-cause mortality, change on 6-point ordinal scale, laboratory biomarkers | hydroxychloroquine, antivirals, antibiotics, interferon beta |
| Mikulska, 2020 | Italy | single centre | Prospective with control | 8mg/kg IV (62%) or 162mg SC (38%). Second dose in 24% | respiratory failure | IMV, death | hydroxychloroquine, antivirals, antibiotics |
| Morena, 2020 | Italy | single centre | Prospective | 8mg/kg IV repeated after 12h | respiratory failure, IL-6 > 40pg/mL | death, hospital discharge | hydroxychloroquine, antivirals, antibiotics |
| Perrone 2020 | Italy | multi-centre | Single arm, open-label & validation | 8mg/kg/IV | respiratory failure | mortality rates at 14 and 30 days | hydroxychloroquine, antibiotics, antivirals, steroids |
| *Rosas, I., 2020 | USA | multi-centre | Placebo-controlled, double blind, phase 3 RCT | 8mg/kg IV, second dose 8-24h later permitted | respiratory failure with bilateral radiological infiltrates | status on a 7-point ordinal scale, time to hospital/ICU discharge, time to improvement on ordinal scale, incidence of IMV | corticosteroids, antivirals, convalescent plasma |
| Roumier, 2020 | France | single centre | Prospective with control | 8mg/kg IV repeated once | respiratory failure, hyperinflammation | mortality, IMV, hospital status | Hydroxychloroquine, azithromycin, corticosteroids |
| Salvarani, 2020 | Italy | multi-centre | Open label RCT | 8mg/kg IV, repeated 12h later | respiratory failure and hyperinflammation | ICU admission and need for IMV, death, respiratory failure | hydroxychloroquine, antivirals, antibiotics |
| *Sanchez-Montalva, 2020 | Spain | single centre | Prospective | 400-600mg IV | respiratory failure, hyperinflammation | death at 7 days, admission to ICU, ARDS | Hydroxychloroquine, antibiotics, antivirals |
| Salama, 2020 | USA | multi-centre | Double blind RCT | 8mg/kg IV | respiratory failure not requiring ventilatory support | mortality, ventilatory requirement, duration of hospitalisation | Antivirals, corticosteroids |
| Sciascia, 2020 | Italy | multi-centre | Prospective | 8mg/kg IV or 324mg S/C. Second dose in 83% | respiratory failure, hyperinflammation | medication safety, oxygen requirement, laboratory biomarkers | antivirals |

| | | | | | | | |
|---|---|---|---|---|---|---|---|
| Stone, 2020 | USA | multi-centre | Double blind RCT | 8mg/kg IV | hyperinflammation with two of: fever, lung infiltrates or respiratory failure | intubation or death, | antiviral, hydroxychloroquine, corticosteroids |
| Strohbehn, 2020 | USA | single centre | Phase 2 open label | 40-200mg | bilateral radiological infiltrates, fever, CRP>40mg/L | resolution of fever, CRP reduction, overall survival at 28 days, rate and duration of IMV, duration of supplemental oxygen | hydroxychloroquine, azithromycin, antiviral |
| Toniati, 2020 | Italy | single centre | Prospective | 8mg/kg IV, repeated after 12h (87%). Third dose 24h later (13%) | respiratory failure requiring ventilatory support | ventilatory requirements, discharge, death | hydroxychloroquine, antivirals, antibiotics, corticosteroids |
| Biran, 2020 | USA | multi-centre | Retrospective | 400mg IV with 12% receiving a second dose | hospitalised requiring ICU stay | mortality, inflammatory biomarkers, oxygenation, infection, use of vasopressors | corticosteroids, hydroxychloroquine, azithromycin |
| Canziani, 2020 | Italy | multi-centre | Retrospective | 8mg/kg IV followed by a second dose 24h later (95%) | respiratory failure, elevated CRP, absence of active bacterial infection | mortality, incidence of invasive ventilation, thromboembolic events, haemorrhagic event, infections | hydroxychloroquine, antivirals, antibiotics, corticosteroids |
| Capra, 2020 | Italy | single centre | Retrospective | 400mg IV (53%); 324mg SC (44%) | tachypnoea or hypoxia. IMV patients excluded | overall mortality | hydroxychloroquine, antivirals |
| Chillmuri, 2020 | USA | single centre | Retrospective | 400mg IV | respiratory failure and hyperinflammation | ventilatory requirement, mortality | hydroxychloroquine, antivirals, corticosteroids |
| De Rossi, 2020 | Italy | single centre | Retrospective | 400mg IV (48%); 324mg SC (52%) | respiratory failure, bilateral radiological infiltrates. IMV patients excluded | overall mortality | hydroxychloroquine, antivirals |
| Eimer, 2020 | Sweden | single centre | Retrospective | 8mg/kg IV | respiratory failure admitted to intensive care, with hyperinflammation | 30-day mortality, time to extubation, ventilator free-days, length of hospital and ICU stay | Nil |
| Fisher, 2020 | USA | single centre | Retrospective | 400mg IV, repeated after 24h | respiratory failure | 30 day mortality | hydroxychloroquine, steroids |

| | | | | | | | |
|---|---|---|---|---|---|---|---|
| Galvan Roman, 2020 | Spain | single centre | Retrospective | 8mg/kg/IV, repeated after 12h | respiratory failure, hyperinflammation, | mortality, IL-6 levels, mechanical ventilation, | hydroxychloroquine, antivirals, antibiotics, corticosteroids |
| *Garcia, 2020 | Spain | single centre | Retrospective | 400-600mg IV repeated 12h apart with up to 3 doses | radiological infiltrates, respiratory failure and hyperinflammation | ICU admission and need for IMV | hydroxychloroquine, antivirals, azithromycin |
| Gokhale, 2020 | India | single centre | Retrospective | 400mg IV | respiratory failure, bilateral radiological infiltrates, hyperinflammation | overall mortality | hydroxychloroquine, antivirals, antibiotics, corticosteroids |
| Guaraldi, 2020 | Italy | multi-centre | Retrospective | 8mg/kg IV, repeated after 12h, or 324mg SC single dose | respiratory failure, lung infiltrates >50% | IMV or death | hydroxychloroquine, antivirals, antibiotics, corticosteroids |
| Guisado-Vasco, 2020 | Spain | single centre | Retrospective | 8mg/kg/IV | radiological infiltrates and respiratory failure | hospital mortality, length of hospitalisation, admission to ICU, requirement for IMV | hydroxychloroquine, antivirals, corticosteroids |
| Gupta, 2020 | USA | multi-centre | Retrospective | Treated in first 2 days, dose not specified | admitted to ICU | hospital mortality, secondary infections | hydroxychloroquine, azithromycin, corticosteroids |
| Hill, 2020 | USA | single centre | Retrospective | 400mg IV, repeated in 3 patients after 24h | fever with either respiratory failure, haemodynamic instability, or serum IL-6 >5 times upper limit of normal | clinical improvement (two-point reduction on six-point scale), mortality within 28 days | hydroxychloroquine, remdesivir |
| Holt, 2020 | USA | single centre | Retrospective | 400mg IV | respiratory failure and hyperinflammation | mortality | N/R |
| Ip, 2020 | USA | multi-centre | Retrospective | 400mg IV | hospitalised on ICU | overall mortality | hydroxychloroquine, azithromycin, corticosteroids |
| Kewan, 2020 | USA | single centre | Retrospective | 8mg/kg IV | respiratory failure, lung infiltrates, hyperinflammation | Time to clinical improvement, duration of IMV, duration of vasopressor support | hydroxychloroquine, azithromycin, corticosteroids |
| Kimmig, 2020 | USA | single centre | Retrospective | 400mg IV | clinical deterioration with hyperinflammation | mortality, infection rate | N/R |
| Klopfenstein, 2020 | France | single centre | Retrospective | N/R | respiratory failure, >25% lung infiltrates, hyperinflammation | death and/or ICU admission | hydroxychloroquine, antivirals, antibiotics, corticosteroids |

| | | | | | | | |
|---|---|---|---|---|---|---|---|
| Lewis, 2020 | USA | multi-centre | Retrospective | 400mg IV | respiratory failure and hyperinflammation | mortality, duration of hospitalisation | azithromycin, corticosteroids |
| Martinez-Sanz, 2020 | Spain | multi-centre | Retrospective | 600-800mg | hospitalised | time to death or intensive care unit admission | hydroxychloroquine, antivirals, antibiotics, corticosteroids |
| # Narain, 2020 | USA | multi-centre | Retrospective | N/R | hyperinflammation | hospital mortality | hydroxychloroquine |
| Nasa, 2020 | India | multi-centre | Retrospective | 8mg/kg IV, repeated after 12 hours | respiratory failure with hyperinflammation | mortality at day 28 | hydroxychloroquine, antivirals, corticosteroids |
| Patel, 2020 | USA | single centre | Retrospective | N/R | severe: respiratory failure critical: requiring IMV | overall mortality, hospital discharge, inflammatory biomarkers | hydroxychloroquine, antivirals, corticosteroids |
| * Petrak, 2020 | USA | multi-centre | Retrospective | N/R | IMV | mortality | hydroxychloroquine, antivirals, antibiotics, corticosteroids |
| Pettit, 2020 | USA | single centre | Retrospective | 400mg IV | respiratory failure with hyperinflammation | infection rate | hydroxychloroquine and remdesivir |
| Potere, 2020 | Italy | single centre | Retrospective | 324mg SC | hyperinflammation with no hypoxaemia | disease progression, inflammatory biomarkers | hydroxychloroquine, antivirals, corticosteroids |
| *Ramaswamy, 2020 | USA | multi-centre | Retrospective | 400mg IV, 8mg/kg | respiratory failure, hyperinflammation | inpatient mortality | hydroxychloroquine, azithromycin, corticosteroids |
| Rodriguez-Bano, 2020 | Spain | multi-centre | Retrospective | N/R | hyperinflammation. IMV patients excluded | intubation, death, secondary bacterial infections, scores on a seven-point ordinal scale | hydroxychloroquine, antivirals, antibiotics, interferon beta |
| Rojas-Marte, 2020 | USA | single centre | Retrospective | N/R | respiratory failure | overall mortality rate | hydroxychloroquine, antivirals, antibiotics, corticosteroids |
| Roomi, 2020 | USA | single centre | Retrospective | N/R | hospitalised | overall mortality, IMV | hydroxychloroquine, corticosteroids |
| Rosas, J., 2020 | Spain | single centre | Retrospective | 400/600mg IV | radiological infiltrates and respiratory failure | admission to ICU, hospital discharge, mortality | hydroxychloroquine, antivirals, antibiotics, corticosteroids |

| | | | | | | | |
|---|---|---|---|---|---|---|---|
| Rossi, 2020 | France | single centre | Retrospective | 400mg IV | respiratory failure. IMV patients excluded | composite of all-cause mortality and invasive ventilation | hydroxychloroquine, antivirals, corticosteroids |
| Rossotti, 2020 | Italy | single centre | Retrospective | 8mg/kg IV repeated 12h later if ongoing fever | respiratory failure, bilateral radiological infiltrates, hyperinflammation | overall survival | hydroxychloroquine, antivirals |
| Ruiz-Antoran, 2020 | Spain | multi-centre | Retrospective | 400-600mg IV repeated up to three doses | respiratory failure, hyperinflammation | in-hospital mortality | hydroxychloroquine, antivirals, antibiotics, corticosteroids |
| Somers, 2020 | USA | single centre | Retrospective | 8mg/kg IV | IMV | survival probability, ordinal scale at day 28 | hydroxychloroquine, corticosteroids |
| Tian, 2020 | China | multi-centre | Retrospective | 4-8mg/kg IV repeated after 12h if ongoing fever | respiratory failure and hyperinflammation | mortality, time from admission to discharge | antivirals, antibiotics, corticosteroids |
| Tsai, 2020 | USA | single centre | Retrospective | 400-800mg IV | respiratory failure and ferritin >300ug/mL | overall mortality | hydroxychloroquine, azithromycin |
| * Wadud, 2020 | USA | single centre | Retrospective | N/R | hospitalised | mortality, discharge, number of days on ventilator, in ICU and in hospital | N/R |
| Zheng, 2020 | China | single centre | Retrospective | 400mg IV, repeat after 24h if persistent fever | severe: respiratory failure critical: shock | mortality, discharge, inflammatory biomarkers | Nil |

**Supplementary Table 1 –** Methodological characteristics of included studies. Age in years reported as mean (standard deviation) unless otherwise stated.
ARDS, acute respiratory distress syndrome; CPAP, continuous positive airways pressure; CRP, C reactive protein; ICU, intensive care unit; IL6, interleukin 6; IV, intravenous; IMV, invasive mechanical ventilation; NIV, non-invasive ventilation; N/R, not reported; SC, subcutaneous; SOFA, sequential organ failure assessment; suPAR, soluble urokinase plasminogen activator receptor. * non peer-reviewed preprint study; #, study investigating both anakinra and tocilizumab

| Author, year | Study design | N Treatment/ Control | Follow up, days | Control Age | Intervention Age | Sex (male control) % | Sex (male) intervention % | Outcomes |
|---|---|---|---|---|---|---|---|---|
| **ANAKINRA** | | | | | | | | |
| Balkhair, 2020 | Prospective with control | 45/24 | N/R | 51.7 (14.8)[a] | 49.8 (16)[a] | 71 | 78 | IMV occurred in 31% in the anakinra group and 75% in the control (p < 0.001). Death occurred in 29% in the anakinra group and 46% in the control (p = 0.082). |
| Huet, 2020 | Prospective with control | 52/44 | N/R | 71 (15)[a] | 71 (13)[a] | 57 | 69 | IMV or death in anakinra group vs control HR 0.22; 95% CI 0.1-0.49. For death alone: HR 0·30; 95% CI 0·12–0·71. Decrease in CRP vs control group. |
| Kooistra, 2020 | Prospective with control | 21/39 | 28 | 67 (59-72)[c] | 63 (55-71)[c] | 85 | 67 | No difference between anakinra and control group in time on IMV (23 vs 17 days; p=0.79), length of ICU stay (24 days vs 17; p=0.59), 28 day mortality (19% vs 18%; p=087) |
| *Kyriazopoulou, 2020 | Prospective | 130/130 | 30 | 63.5 (13.7) | 63.2 (14.1) | 65 | 62 | severe respiratory failure lower in anakinra treated group (22.3% vs 59.2%), and lower 30-day mortality (aHR 0.49, 95%CI 0.25-0.97). |
| Cauchois, 2020 | Retrospective | 12/10 | N/R | N/R | N/R | N/R | N/R | Fewer no. days with oxygen < 3L/min in anakinra group vs control at day 20 (p<0.05). No. of days without IMV similar. Rapid reduction of CRP with anakinra vs. controls (p<0.001) |
| Cavalli, 2020 | Retrospective | 29/16 | 21 | 70 (64-78)[c] | 63 (51-73)[c] | 88 | 83 | Control: Survival at 21 days of 56%. Mechanical ventilation-free survival 50%. Tocilizumab high dose: Survival of 90% at 21 day (p=0.009 vs control group). IMV-free survival 72% (p=0.15 vs control group) |
| # Narain, 2020 | Retrospective | 57/3076 | N/R | 65 (54-77)[c] | 67 (58-75)[c] | 62 | 67 | No effect on mortality (aHR 0.79; 95% CI 0.44-1.42) |
| **SARILUMAB** | | | | | | | | |
| Benucci, 2020 | Prospective | 8/0 | 14 | - | 62 | - | 75 | 87% discharged within 14 days. |

| | | | | | | | | |
|---|---|---|---|---|---|---|---|---|
| Della-Torre, 2020 | Prospective with control | 28/28 | 28 | 57 (52-60) [c] | 56 (49-60) [c] | 71 | 85 | Survival similar in both groups (HR 0.36; 95% CI 0.08-1.68). In treatment group, median time to death higher (19 vs. 4 days; p=0.006), median time to CRP normalisation lower (6 vs. 12 days; p<0.0001). Median time to clinical improvement, discharge and IMV free survival similar. Median time to clinical improvement shorter in patients with a baseline PaO2/FiO2 >100mgHg (7 vs 28 days; HR 0.18; 95% CI 0.02-0.26) |
| * Gordon, 2021 | Adaptive RCT | 45/397 | NR | 61.1 (12.8) [a] | 63.4 (13.4) [a] | 70 | 81 | Mean adjusted odds ratio for survival was 2.01 (95%CI 1.18-2.71). Compared with control, median adjusted odds ratios for organ support-free days was 1.76 (95%CI 1.17-2.91). Sarilumab associated with improved time to ICU discharge (aHR 1.64; 95%CI 1.21-2.45), improved time to hospital discharge (aHR 1.6; 95%CI 1.17-2.40), improved ordinal scale outcomes at day 14 (aOR 1.86; 95%CI 1.22-2.91). |
| Gremese, 2020 | Prospective | 53/0 | 16 (14-24) [b] | - | 66 (40-95) [c] | - | 89 | 83% (89.7% in medical wards and 64.3% in ICU) improved on therapy. Overall mortality of 5.7% |
| Sinha, 2020 | Prospective | 255/0 | N/R | - | 59 (47-70) [c] | - | 63 | 10.9% of patients died. Mortality was lower in patients with FiO2 < 0.45 (HR 0.24; 95% CI 0.08-0.74) |
| **SILTUXIMAB** | | | | | | | | |
| * Gritti 2020 | Prospective with control | 30/30 | 33.3 (7-58) [b] | 65 (56-70) [b] | 64 (57-66) [b] | 80 | 77 | 30-day mortality lower in treatment arm (HR 0.46; 95% CI 0.22-0.97). 53% recovered and were discharged. |
| **TOCILIZUMAB** | | | | | | | | |
| Albertini, 2020 | Prospective with control | 22/22 | 14 | 65 (41-82) [b] | 64 (41-80) [b] | 68 | 73 | average respiratory rate at d14 lower in treated (21.5 vs 25.5 breaths/min; 95% CI -7.5 to -0.4). No difference in requirement for intubation. Significant fall in CRP in treated patients on d7 (p=0.04) |
| Antony, 2020 | Prospective | 80/0 | N/R | - | 63 (51-72) [b] | - | 57 | 8.8% of patients died and 11.3% required mechanical ventilation. CRP levels reduced post therapy, whereas IL-6 increased |

| | | | | | | | | |
|---|---|---|---|---|---|---|---|---|
| Campins, 2020 | Prospective | 58/0 | N/R | - | 60.6 | - | 72 | 32.4% of patients were admitted to intensive care, 13.8% died. No difference in median CRP and IL-6 between survivors and dead |
| * Carvalho, 2020 | Prospective with control | 29/24 | 14 | 59 (51-72) [c] | 55 (44-65) [c] | 75 | 62 | Tocilizumab not associated with mortality (HR 3.97; 95% CI 0.28-5.72), or positive cultures (OR 1.73; 95% CI 0.22-13.82) |
| Dastan, 2020 | Prospective | 42/0 | 28 | - | 56 (44-61) [c] | - | 64 | 14% required IMV, remaining patients showed clinical improvement. By d28, 16.7% of patients died |
| * Gordon, 2021 | Adaptive RCT | 350/397 | NR | 61.1 (12.8) [a] | 61.5 (12.5) [a] | 70 | 74 | Mean adjusted odds ratio for survival was 1.64 (95%CI 1.14-2.35). Compared with control, median adjusted odds ratios for organ support-free days was 1.64 (95%CI 1.25-2.14). Tocilizumab associated with improved time to ICU discharge (aHR 1.42; 95%CI 1.18-1.70), improved time to hospital discharge (aHR 1.41; 95%CI 1.18-1.70), improved ordinal scale outcomes at day 14 (aOR 1.83; 95%CI 1.40-2.41). |
| Hermine, 2020 | Open label RCT | 64/67 | 90 | 63 (57-72) [c] | 64 (57-74) [c] | 66 | 70 | At day 14, fewer patients died or needed ventilation compared with controls (aHR 0.58; 90% CI 0.30-1.09). At day 28, mortality was similar in both groups (aHR 0.92; 95%CI 0.33-2.53) |
| Malekzadeh, 2020 | Prospective | 126/0 | 14 | - | 54 (13) [a] | - | 64 | By day 14, 4.7% (4/86) of severe patients and 50% (20/40) of critical patients died. By the end, 7% (6/86) of severe patients and 60% (24/40) of critical patients died. |
| Mikulska, 2020 | Prospective with control | 29/66 | 53 (4-70) [b] | 68 (13) [a] | 66 (10) [a] | 67 | 83 | 14-day mortality was 13.8% vs. 21.8% in control group. Mortality at study end lower in treatment group (HR 0.48; 95% CI 0.23-0.99) |
| Morena, 2020 | Prospective | 51/0 | 30 | N/A | 60 (50-70) [c] | N/A | 78 | Over a median follow up of 34 days, 67% of patients showed an improvement in clinical severity. Overall mortality rate was 27% |
| Perrone, 2020 | Single-arm, open-label phase 2 trial | 180/121 | 30 | ≤60: 36%<br>61-70: 33%<br>≥71: 31% | ≤60: 44%<br>61-70: 37%<br>≥71: 19% | 77 | 83 | Pre-specified expected lethality rates defined as 20% and 35% at 14 and 30 days respectively. Lethality rates were 18.4% (95% CI 13.6-24.0, p=0.52) and 22.4% (95% CI 17.2-28.3, p<0.001) at 14 and 30 days. In tocilizumab group alone, lethality rates were 15.6% and 20%. |
| Perrone, 2020 | Prospective with control | 528/360 | 30 | ≤60: 43%<br>61-70: 30%<br>≥71: 27% | ≤60: 40%<br>61-70: 28%<br>≥71: 32% | 77 | 83 | In the validation cohort, lethality rates were consistently lower than the predefined null hypothesis both at 14 and 30 days in the overall cohort (11.4% and 18.4%) and in the tocilizumab only group (10.9% and 20.0%) |

| | | | | | | | | |
|---|---|---|---|---|---|---|---|---|
| * Rosas, I., 2020 | Placebo-controlled, double phase 3 RCT | 294/144 | 60 | 61 (14) [a] | 61 | 70 | 70 | No improvement in clinical status at day 28 (p=0.36), or mortality. Ordinal scale values similar (OR 1.19; 95% CI 0.81-1.76). Median time to hospital discharge shorter with tocilizumab than placebo (20 and 28 days; HR 1.35 95% CI 1.02-1.79). Median duration of ICU stay shorter with tocilizumab (9.8 and 15.5 respectively, p=0.045). Median time to improvement from baseline in 2 or more categories on ordinal scale was 14 days (12-17) in tocilizumab arm and 18 (15-28) days in placebo (p=0.08). Incidence of IMV was 27.9% in tocilizumab arm and 36.7% in placebo (p=0.14) |
| Roumier, 2020 | Prospective with control | 49/47 | 28 | 62 (13) [a] | 58 (12) [a] | 81 | 82 | Tocilizumab reduced requirement for IMV (aHR 0.58; 95% CI 0.36-0.94). No difference in mortality (aHR 0.68; 95% CI 0.31-1.75) |
| Salama, 2020 | Double-blind RCT | 249/128 | 60 | 55.6 (14.9)[a] | 56 (14.3) [a] | 57 | 60 | IMV or death at day 28 was lower in tocilizumab group (aHR 0.56; 95% CI 0.33 - 0.97). Mortality similar in both groups (10.4% vs 8.6%). |
| Salvarani, 2020 | Open label RCT | 60/63 | 30 | 60 (54-69) [c] | 62 (52-74) [c] | 56 | 67 | 28% in the tocilizumab arm and 27% in SOC group showed clinical worsening within 14 days (RR, 1.05; 95% CI, 0.59-1.86). Mortality at 14 days and at 30 days (was comparable in the 2 groups |
| * Sanchez-Montalva, 2020 | Prospective | 82/0 | N/R | - | 59 (20) [a] | - | 63 | Mortality at 7 days was 26.8%. ARDS developed in 54.9% |
| Sciascia, 2020 | Prospective | 63/0 | 14 | - | 63 (13) [a] | - | 88 | Tocilizumab associated with increased survival (HR 2.2; 95% CI 1.3-6.7). Overall mortality was 11% |
| Stone, 2020 | Double blind RCT | 161/82 | 28 | 57 (45-70) [c] | 62 (46-70) [c] | 55 | 60 | HR for intubation or death compared with placebo was 0.83;95% CI, 0.38 to 1.81. At 14 days, 18.0% in tocilizumab and 14.9% in of placebo had disease progression. At 14 days, 24.6% of tocilizumab group and 21.2% of placebo were receiving supplemental oxygen. |
| Strohbehn, 2020 | Phase 2 open label trial with control | 32/41 | 28 | 68 (58-78) [c] | 69 (41-73) [c] | 59 | 50 | At 24 hours, 75% of tocilizumab vs 34.1% of control were afebrile (p=0.001). 86.2% of tocilizumab vs. 14.3% control achieved CRP decrease of at least 25% (p<0.001). Median time to recovery was 3 days (IQR 2-5) |

| | | | | | | | | |
|---|---|---|---|---|---|---|---|---|
| Toniati, 2020 | Prospective | 100/0 | 10 | - | 62 (57-71) [c] | - | 88 | Overall at 10 days 77% of patients improved or stabilised and 23% worsened. Mortality was 20% |
| Biran, 2020 | Retrospective | 210/420 | 22 (11-53) [c] | 65 (56-74) [c] | 62 (53-71) [c] | 67 | 74 | Exposure to tocilizumab was associated with lower hospital mortality (HR 0.64; 95% CI 0.47-0.87). In subgroup analyses, tocilizumab associated with decreased hospital mortality in those with a CRP≥150mg/L (HR 0.48;95% CI 0.3-0.77), but not in those with CRP>150mg/L (HR 0.92;95% CI 0.57-1.48). |
| Canziani, 2020 | Retrospective | 64/64 | N/R | 64 (8) [a] | 63 (12) [a] | 73 | 73 | 30-day mortality unaffected (aHR 0.82; 95% CI 0.42-1.58). Between days 6 and 30, HR 0.41 (95% CI 0.17-0.96) for tocilizumab vs controls. Tocilizumab associated with lower risk of IMV (HR 0.36; 95% CI 0.16-0.83). No effect on thrombotic events, bleeding, infection |
| Capra, 2020 | Retrospective | 62/23 | 28 | 70 (55-80) [c] | 63 (54-73) [c] | 83 | 73 | Tocilizumab associated with reduced risk of mortality (HR 0.035; 95% CI 0.004-0.347) |
| Chillmuri, 2020 | Retrospective | 83/685 | N/R | 63 (54-73) [c] | 60 (50-70) [c] | 61 | 74 | Tocilizumab associated with lower composite endpoint of IMV or death (aHR 0.29; 95% CI 0.16-0.54) |
| De Rossi, 2020 | Retrospective | 90/68 | N/R | 71 (15) [a] | 63 (13) [a] | 72 | 71 | Tocilizumab group associated with reduced risk of mortality (aHR 0.057; 95% CI 0.017-0.187). Survival rate or mean time to discharge did not differ between two administration (IV and SC) routes. |
| Eimer, 2020 | Retrospective | 22/22 | 30 | 60 (54-67) [c] | 61 (49-64) [c] | 77 | 96 | No difference in all-cause mortality at 30 days (HR 0.52; 95% CI 0.19-1.39).Median time to death was 8 days in treated (IQR 5-12.5) and 14 days (IQR 10-19, p = 0.15) in control. In tocilizumab group, significantly more ventilator free days. Freedom from IMV was achieved earlier and in a higher proportion of patients (HR 2.83; 95% CI 1.48-5.4). Length of hospital stay shorter in tocilizumab group |
| Fisher, 2020 | Retrospective | 45/70 | 30 | 60.6 (13.4)[a] | 56.2 (14.7) [a] | 73 | 64 | No difference in mortality associated with tocilizumab (OR 1.04, 95% C.I. 0.27 – 3.75) |
| Galvan Roman, 2020 | Retrospective | 58/88 | 61 (58-64) [c] | 64 (54-72) [b] | 61 (54-70) [c] | 65 | 69 | patients with high IL-6 not treated with TCZ showed high 139 mortality (HR: 4.6; p=0.003), as well as those with low IL-6 treated with tocilizumab (HR: 3.6; p=0.016). |

| | | | | | | | | |
|---|---|---|---|---|---|---|---|---|
| * Garcia, 2020 | Retrospective | 77/94 | 14.7 (10.6) [a] | 61 (16) [a] | 62 (12) [a] | 63 | 69 | Tocilizumab associated with fewer ICU admissions (10.3% vs. 27.6%; p=0.005) and need for IMV (0 vs 13.8%, OR 0.03, 95% CI 0.007-0.1) |
| Gokhale, 2020 | Retrospective | 70/91 | 31 (12-48) [c] | 55 (48-65) [c] | 52 (44-57) [c] | 58 | 67 | Tocilizumab associated with reduced mortality (HR 0.616;95% CI 0.38-0.99) |
| Guaraldi, 2020 | Retrospective | 179/365 | 12 (6-17) [c] | 69 (57-78) [c] | 64 (54-72) [c] | 64 | 71 | Tocilizumab use associated with reduced risk of death (7% vs. 20%; aHR 0.38; 95% CI 0.17-0.83) and composite outcome of IMV or death (aHR 0.61;95% CI 0.4-0.92). |
| Guisado-Vasco, 2020 | Retrospective | 132/475 | N/R | N/R | 69 (22) [c] | N/R | 65 | Increased mortality with tocilizumab (aOR 2·4, 95% CI, 1·13 - 5·11) |
| Gupta, 2020 | Retrospective | 433/3492 | 26 (15-38) [c] | 63 (52-72) [c] | 58 (48-65) [c] | 62 | 69 | Patients treated with tocilizumab had a lower risk of death compared with those not treated with tocilizumab (HR, 0.71; 95% CI, 0.56-0.92) |
| Hill, 2020 | Retrospective | 43/45 | 28 | N/R | N/R | 69 | 70 | Tocilizumab not associated with lower risk of mortality (aHR 0.57; 95% CI 0.21-1.52) or a difference in clinical improvement (aHR 0.92; 95% CI 0.38-2.22) |
| Holt, 2020 | Retrospective | 24/30 | N/R | N/R | N/R | N/R | N/R | In multivariate analysis, tocilizumab administration had no effect on mortality (OR 0.32; 95% CI 0.02-3.69) |
| Ip, 2020 | Retrospective | 134/413 | N/R | 69 (58-77) [c] | 62 (533-70) [c] | 62 | 74 | Tocilizumab associated with reduced mortality within the ICU setting (aHR 0.76; 95% CI 0.57-1.00) |
| Kewan, 2020 | Retrospective | 28/23 | 10 (6-17) [c] | 70 (55-75) [c] | 62 (53-71) [c] | 48 | 71 | Median time to clinical improvement in tocilizumab vs. no tocilizumab was 6.5 days (IQR 4-9) vs. 7 days (IQR 5-10) among all patients (HR 1.14; 95% CI 0.55-2.38). Shorter median length of hospital stay with tocilizumab. The median duration of vasopressor support and IMV were 2 days (IQR: 1·75 – 4·25 days) vs. 5 days (IQR: 4 – 8 days), p = 0.039, and 7 days (IQR: 4 – 14 days) vs. 10 days (IQR: 5 – 15 days) in tocilizumab vs. no tocilizumab cohorts, p = 0.11 |
| Kimmig, 2020 | Retrospective | 54/57 | N/R | 62 (17) [a] | 65 (14) [a] | 44 | 69 | Tocilizumab was associated with higher risk of mortality (35.2% vs 19.3%, p=0.02) |

| | | | | | | | | |
|---|---|---|---|---|---|---|---|---|
| Klopfenstein, 2020 | Retrospective | 20/25 | N/R | 71 (15) [a] | 77 (11) [a] | N/R | N/R | Death and/or ICU admissions higher in tocilizumab cohort vs control (72% vs 25%; p=0.002). No difference in death alone (25% vs 48%; p=0.0066) |
| Lewis, 2020 | Retrospective | 497/497 | N/R | 64 (52-76) [c] | 61 (52-69) [c] | 58 | 71 | Tocilizumab associated with improved survival (aHR 0.24; 95% CI 0.18-0.33). Similar time to hospital discharge (aHR 0.86; 95% CI 0.78-1.17) |
| Martinez-Sanz, 2020 | Retrospective | 260/969 | 6 (3-9) [c] | 68 (57-80) [c] | 65 (55-76) [c] | 59 | 73 | In patients with CRP>150mg/L, tocilizumab associated with decreased risk of death (aHR 0.34; 95% CI 0.16-0.72) and ICU admission or death (aHR 0.38; 95% CI 0.19-0.81), but not in those with CRP <150mg/L. For all patients, tocilizumab not associated with risk of death (HR 0.77; 95% CI 0.48-1.22) |
| # Narain, 2020 | Retrospective | 73/3076 | N/R | 65 (54-77) [c] | 62 (55-69) [c] | 65 | 71 | No effect on mortality (aHR 0.79; 95% CI 0.47-1.32) |
| Nasa, 2020 | Retrospective | 22/63 | N/R | 52 [a] | 51 [a] | 95 | 100 | mortality at day 7 and 28 was significantly lower in the tocilizumab group (p = 0.007 and p = 0.001 respectively). |
| Patel, 2020 | Retrospective | 42/41 | 19 (5.5) [c] | 67 (20-91) [b] | 68 (25-96) [b] | 49 | 50 | CRP improved in all tocilizumab patients. No difference in mortality with tocilizumab but more patients discharged compared with controls (55% vs 24%) |
| * Petrak, 2020 | Retrospective | 81/37 | N/R | 62.3 (12.9)[a] | 56.3 (12.7) [a] | 57 | 67 | No difference between tocilizumab and mortality (aOR 0.83; 95%CI 0.34-1.98). However early therapy was associated with reduced mortality (aOR 0.15; 95%CI 0.04-0.5) |
| Pettit, 2020 | Retrospective | 74/74 | 58 | 65 (16) [a] | 66 (14) [a] | 45 | 58 | Mortality rate higher in tocilizumab cohort (39% vs 23%; p=0.03). |
| Potere, 2020 | Retrospective | 10/10 | N/R | 56 (49-60) [c] | 55 (54-60) [c] | 60 | 60 | Tocilizumab associated with reduction in CRP over three days. None of the tocilizumab patients had disease progression (requirement of oxygen or mechanical ventilation) whereas progression occurred in 50% of control group |
| *Ramaswamy, 2020 | Retrospective | 21/65 | N/R | 64 (16) [a] | 63 (16) [a] | 55 | 62 | Mortality lower in tocilizumab group (HR 0.25; 95% CI 0.07-0.9) |

| | | | | | | | | |
|---|---|---|---|---|---|---|---|---|
| Rodriguez-Bano, 2020 | Retrospective | 88/344 | 21 | 69 (59-76) [c] | 66 (56-72) [c] | 69 | 72 | Tocilizumab associated with reduced risk of death (aHR 0.12; 95% CI 0.02-0.56) and reduced risk of composite outcome of intubation or death (aHR 0.32; 95% CI 0.15-0.67) |
| Rojas-Marte, 2020 | Retrospective | 96/97 | 14.5 (8.8) [a] | 62 (14) [a] | 58 (14) [a] | [a]65 | 77 | Similar mortality in both groups (52% vs 61%; p=0.09) |
| Roomi, 2020 | Retrospective | 32/144 | N/R | 66 | 58 | 45 | 64 | No difference in hospital mortality (aOR 0.28; 95% CI 0.05-1.4), IMV (aOR 1.2;95% CI 0.49-2.9) and hospital discharge (aOR 0.78;95% CI 0.28-2.1). Reduction in CRP levels on day 7 compared with control (21% vs 56%; OR 0.21; 95% CI 0.08-0.55 |
| Rosas, J., 2020 | Retrospective | 20/17 | 30 | 73.8 (14.8)[a] | 59.4 (14.5) [a] | 65 | 75 | Mortality was 20% in tocilizumab group and 35% in control group. Admission to ICU was 65% in tocilizumab and 0% in control |
| Rossi, 2020 | Retrospective | 84/84 | 28 | 64 (17) [a] | 65 (13) [a] | 58 | 66 | Tocilizumab associated with reduced mortality (aHR 0.42; 95% CI 0.22-0.82), and reduced composite of mortality or IMV (aHR 0.34; 95% CI 0.22-0.52) |
| Rossotti, 2020 | Retrospective | 74/148 | N/R | 59 (52-70) [c] | 59 (51-71) [c] | 81 | 82 | Tocilizumab associated with reduced mortality (unadjusted HR 0.49; 95% CI 0.26-0.95), but longer hospital stay (HR 1.66; 95% CI 1.09-2.52) |
| Ruiz-Antoran, 2020 | Retrospective | 268/238 | 12 (7-18) [b] | 71 14) [a] | 65 (12) [a] | 59 | 69 | Mortality lower in patients treated with tocilizumab than controls (16.8% vs. 31.5%, aHR 0.74; 95%CI 0.62-0.89) |
| Somers, 2020 | Retrospective | 78/76 | N/R | 60 (15) [a] | 55 (15) [a] | 64 | 68 | Tocilizumab associated with lower risk of death (aHR 0.55; 95% CI 0.33-0.9) |
| Tian, 2020 | Retrospective | 65/130 | NR | 67.5 (61-75) [c] | 71(63-75) [c] | 63 | 74 | Mortality lower in tocilizumab group (aHR 0.47; 95%CI 0.25-0.9) |
| Tsai, 2020 | Retrospective | 66/66 | N/R | 61 (16) [a] | 62 (14) [a] | 76 | 70 | No difference in mortality between two groups (OR 1.0;95% CI 0.465-2.151) |
| * Wadud, 2020 | Retrospective | 44/50 | N/R | 66 [b] | 56 [b] | 70 | 84 | Lower mortality in tocilizumab group (38.6% vs. 52%; p<0.001) |

| | | | | | | | | |
|---|---|---|---|---|---|---|---|---|
| Zheng, 2020 | Retrospective | 92/89 | 28 (6-62) [b] | 67 (25-85) [b] | 69 (25-87) [b] | 53 | 62 | Increased mortality in tocilizumab group, but significant reduction in CRP level at 1 week |

**Supplementary Table 2** – Patient characteristics and outcomes of included studies. Absolute numbers reported for follow up days unless otherwise statement. Number of males in control and intervention group reported as percentage (%)

[a], mean and standard deviation; [b], median and range; [c], median and interquartile range; aHR, adjusted hazard ratio; CI, confidence interval; CRP, C-reactive protein; ICU, intensive care unit; IL6, interleukin-6; IMV, invasive mechanical ventilation; IV, intravenous; N/R, not reported; OR, odds ratio; SC, subcutaneous; -, not available; * non peer-reviewed preprint study #, study investigating both anakinra and tocilizumab

| Randomised controlled trials | | | | | | |
|---|---|---|---|---|---|---|
| **Tocilizumab** | | | | | | |
| | Gordon 2021 * | Hermine 2020 | Rosas, I. 2020 * | Salama 2020 | Salvarani 2020 | Stone 2020 |
| **Randomisation** | Low | Low | Low | Low | Low | Low |
| **Intervention assignment** | Low | High | Low | Low | High | Low |
| **Intervention adherence** | Low | Some concern | Low | Low | Some concern | Low |
| **Missing data** | Some concern | Low | Low | Low | Low | Low |
| **Outcome** | Low | Low | Low | Low | Low | Low |
| **Results** | Low | Low | Low | Low | Low | Low |
| **Overall risk of bias** | Low | Some concern | Low | Low | Some concern | Low |

**Supplementary Table 3(a) –** Risk of bias assessment for randomised clinical trials using Cochrane risk of bias 2 tool. Risk of bias was assessed in six categories and scored as either low risk of bias, some concern, or high risk of bias, before an overall risk of bias was given to each study.

* non peer-reviewed preprint study

**Prospective studies**

**Tocilizumab**

| | Albertini 2020 | Antony 2020 | Campins 2020 | Carvalho 2020 * | Dastan 2020 | Malekzadeh, 2020 | Mikulsa 2020 | Morena 2020 | Perrone 2020 | Roumier, 2020 | Sanchez-Motalva 2020 * | Sciascia 2020 | Strohbehn 2020 | Toniati 2020 |
|---|---|---|---|---|---|---|---|---|---|---|---|---|---|---|
| 1 | + | + | - | + | + | + | + | + | + | + | + | - | + | + |
| 2 | + | + | - | + | + | + | + | + | + | + | + | + | + | + |
| 3 | + | + | CD | CD | + | CD | + | + | + | + | + | CD | + | + |
| 4 | + | + | + | + | + | + | + | + | + | + | + | + | + | + |
| 5 | - | - | - | - | - | - | - | - | + | + | - | - | - | - |
| 6 | + | + | + | + | + | + | + | + | + | + | + | + | + | + |
| 7 | + | - | CD | + | + | + | + | + | + | + | + | + | + | + |
| 8 | N/A | N/A | N/A | N/A | N/A | N/A | N/A | N/A | N/A | N/A | N/A | N/A | N/A | N/A |
| 9 | - | + | - | - | + | + | - | + | + | + | - | - | + | - |
| 10 | N/A | N/A | N/A | N/A | N/A | N/A | N/A | N/A | N/A | N/A | N/A | N/A | N/A | N/A |
| 11 | + | - | - | + | + | + | + | + | + | + | + | + | + | - |
| 12 | - | - | - | - | - | - | - | - | - | - | - | - | - | - |
| 13 | + | + | CD | + | + | + | + | + | + | + | + | + | + | + |
| 14 | - | - | - | + | - | - | + | + | - | + | + | - | + | - |
| Total | 8 | 7 | 2 | 8 | 9 | 8 | 9 | 10 | 10 | 11 | 9 | 6 | 10 | 7 |
| Rating | **Fair** | **Fair** | **Poor** | **Fair** | **Fair** | **Fair** | **Fair** | **Good** | **Good** | **Good** | **Fair** | **Poor** | **Good** | **Fair** |

| Prospective studies | | | | | | | | | |
|---|---|---|---|---|---|---|---|---|---|
| | **Anakinra** | | | | **Sarilumab** | | | | **Siltuximab** |
| | Balkhair, 2020 | Huet 2020 | Kooistra, 2020 | Kyriazopoulou, 2020 * | Benucci 2020 | Della-Torre 2020 | Sinha 2020 | Gremese 2020 | Gritti 2020 * |
| 1 | + | + | + | + | + | + | + | + | + |
| 2 | + | + | + | + | - | + | + | + | + |
| 3 | + | + | + | + | CD | + | + | + | + |
| 4 | + | + | - | + | - | + | + | + | + |
| 5 | + | + | - | + | - | - | - | - | - |
| 6 | + | + | + | + | + | + | + | + | + |
| 7 | + | CD | + | + | + | + | + | + | + |
| 8 | N/A | N/A | N/A | N/A | N/A | N/A | N/A | N/A | N/A |
| 9 | + | + | + | + | + | + | + | + | + |
| 10 | N/A | N/A | N/A | N/A | N/A | N/A | N/A | N/A | N/A |
| 11 | + | + | + | + | + | + | + | + | + |
| 12 | - | - | - | - | - | - | - | - | - |
| 13 | + | + | + | + | + | + | + | + | + |
| 14 | - | + | - | + | - | + | + | - | + |
| Total | 10 | 10 | 8 | 10 | 6 | 10 | 10 | 9 | 10 |
| Rating | **Good** | **Good** | **Fair** | **Good** | **Poor** | **Good** | **Good** | **Fair** | **Good** |

**Supplementary Table 3(b).** Risk of bias assessment for prospective studies. Questions numbered in the first column. 1. Was the research question or objective in this paper clearly stated? 2. Was the study population clearly specified and defined? 3. Was the participation rate of eligible persons at least 50%? 4. Were all the subjects selected or recruited from the same or similar populations (including the same time period)? Were inclusion and exclusion criteria for being in the study prespecified and applied uniformly to all participants? 5. Was a sample size justification, power description, or variance and effect estimates provided? 6. For the analyses in this paper, were the exposure(s) of interest measured prior to the outcome(s) being measured? 7. Was the timeframe sufficient so that one could reasonably expect to see an association between exposure and outcome if it existed? 8. For exposures that can vary in amount or level, did the study examine different levels of the exposure as related to the outcome (e.g., categories of exposure, or exposure measured as continuous variable)? 9. Were the exposure measures (independent variables) clearly defined, valid, reliable, and implemented consistently across all study participants? 10. Was the exposure(s) assessed more than once over time? 11. Were the outcome measures (dependent variables) clearly defined, valid, reliable, and implemented consistently across all study participants? 12. Were the outcome assessors blinded to the exposure status of participants? 13. Was loss to follow-up after baseline 20% or less? 14. Were key potential confounding variables measured and adjusted statistically for their impact on the relationship between exposure(s) and outcome(s)?

+, criteria satisfied; - , not satisfied; N/A, not applicable; CD, cannot determine; * non peer-reviewed preprint study

**Retrospective studies**

**Tocilizumab**

| | Biran 2020 | Canziani 2020 | Capra 2020 | Chillmuri, 2020 | De Rossi 2020 | Eimer 2020 | Fisher, 2020 | Galvan-Roman 2020 | Garcia 2020 * | Gokhale 2020 | Guaraldi 2020 | Guisado-Vasco 2020 | Gupta 2020 | Hill 2020 | Holt 2020 | Ip 2020 | Kewan 2020 | Kimmig 2020 |
|---|---|---|---|---|---|---|---|---|---|---|---|---|---|---|---|---|---|---|
| 1 | + | + | + | + | + | + | + | + | + | - | + | + | + | + | + | + | + | + |
| 2 | + | + | + | + | + | - | + | + | + | + | + | + | + | + | + | + | + | + |
| 3 | - | - | + | - | + | - | - | - | - | - | - | - | - | - | - | - | - | - |
| 4 | + | + | + | + | + | + | + | + | + | + | + | + | + | + | CD | + | + | + |
| 5 | + | + | + | - | + | - | + | CD | - | + | - | + | + | - | CD | + | - | CD |
| 6 | + | + | + | + | + | + | + | + | + | + | + | - | + | + | + | + | + | + |
| 7 | + | + | + | + | + | + | CD | CD | + | + | + | + | + | + | CD | + | + | + |
| 8 | - | - | - | - | - | - | - | - | - | - | - | - | - | - | - | - | - | - |
| 9 | + | + | + | + | + | + | + | + | + | + | + | + | + | + | + | + | + | + |
| 10 | - | - | - | - | - | + | + | + | + | + | - | - | + | - | + | + | + | - |
| 11 | - | - | - | - | - | - | - | - | - | - | - | - | - | - | - | - | - | - |
| 12 | + | + | + | + | + | + | + | - | - | + | + | + | + | + | + | + | + | + |
| **Total** | 8 | 8 | 9 | 7 | 9 | 7 | 8 | 6 | 7 | 8 | 7 | 7 | 9 | 7 | 6 | 9 | 8 | 7 |
| **Rating** | **Fair** | **Fair** | **Good** | **Fair** | **Good** | **Fair** | **Fair** | **Fair** | **Fair** | **Fair** | **Fair** | **Fair** | **Good** | **Fair** | **Fair** | **Good** | **Fair** | **Fair** |

**Retrospective studies**

**Tocilizumab**

| | Klopfenstein 2020 | Lewis, 2020 | Martinez-Sanz 2020 | Narain 2020 | Nasa 2020 | Patel 2020 | Petrak 2020 * | Pettit 2020 | Potere 2020 | Ramaswamy 2020 * | Rodriguez-Bano 2020 | Rojas-Marte 2020 | Roomi 2020 | Rosas, J.2000 | Rossi 2020 | Rossotti 2020 | Ruiz-Antoran 2020 | Somers 2020 |
|---|---|---|---|---|---|---|---|---|---|---|---|---|---|---|---|---|---|---|
| 1 | + | + | + | + | + | + | + | + | + | + | + | + | + | + | + | + | + | + |
| 2 | + | + | + | + | + | - | + | + | + | + | + | + | + | + | + | + | + | + |
| 3 | - | - | - | - | - | - | - | - | - | - | - | - | - | - | - | - | - | - |
| 4 | + | + | + | + | - | + | + | + | + | + | + | + | CD | CD | + | + | + | + |
| 5 | - | + | - | + | - | - | + | + | + | + | + | CD | - | - | + | + | CD | CD |
| 6 | + | + | + | + | + | + | + | + | + | + | + | + | + | + | + | + | + | + |
| 7 | + | + | + | + | + | + | + | + | + | + | + | CD | CD | CD | + | + | + | CD |
| 8 | - | - | - | - | - | - | - | - | - | - | - | - | - | - | - | - | - | - |
| 9 | + | + | + | + | + | + | + | + | + | + | + | + | + | + | + | + | + | + |
| 10 | - | + | - | - | - | CD | - | - | + | + | CD | CD | - | + | + | + | - | + |
| 11 | - | - | - | - | - | - | - | - | - | - | - | - | - | - | - | - | - | - |
| 12 | - | + | + | + | - | - | + | - | - | + | + | - | + | - | + | + | + | + |
| **Total** | 6 | 9 | 7 | 8 | 5 | 5 | 8 | 7 | 8 | 9 | 8 | 4 | 4 | 5 | 9 | 9 | 7 | 7 |
| **Rating** | **Poor** | **Good** | **Fair** | **Fair** | **Poor** | **Poor** | **Fair** | **Fair** | **Fair** | **Good** | **Fair** | **Poor** | **Poor** | **Poor** | **Good** | **Good** | **Fair** | **Fair** |

| Retrospective studies | | | | | | | |
|---|---|---|---|---|---|---|---|
| | Tocilizumab | | | | Anakinra | | |
| | Tian 2020 | Tsai 2020 | Wadud 2020 * | Zheng 2020 | Cauchois 2020 | Cavalli 2020 | Narain 2020 |
| 1 | + | + | - | + | + | + | + |
| 2 | + | + | - | + | + | + | + |
| 3 | - | - | - | - | - | - | - |
| 4 | + | + | + | CD | + | + | + |
| 5 | + | + | - | - | + | + | + |
| 6 | + | + | + | + | + | + | + |
| 7 | + | + | CD | CD | + | + | + |
| 8 | - | - | - | - | - | - | - |
| 9 | + | + | + | + | + | + | + |
| 10 | + | + | - | + | + | + | - |
| 11 | - | - | - | - | - | - | - |
| 12 | + | + | - | - | - | - | + |
| **Total** | 9 | 9 | 3 | 5 | 8 | 8 | 8 |
| **Rating** | **Good** | **Good** | **Poor** | **Poor** | **Fair** | **Fair** | **Fair** |

**Supplementary Table 3(c).** Risk of bias assessment for Retrospective studies. 1. Was the research question or objective in this paper clearly stated and appropriate? 2. Was the study population clearly specified and defined? 3. Did the authors include a sample size justification? 4. Were controls selected or recruited from the same or similar population that gave rise to the cases (including the same timeframe)? 5. Were the definitions, inclusion and exclusion criteria, algorithms or processes used to identify or select cases and controls valid, reliable, and implemented consistently across all study participants? 6. Were the cases clearly defined and differentiated from controls? 7. If less than 100 percent of eligible cases and/or controls were selected for the study, were the cases and/or controls randomly selected from those eligible? 8. Was there use of concurrent controls? 9. Were the investigators able to confirm that the exposure/risk occurred prior to the development of the condition or event that defined a participant as a case? 10. Were the measures of exposure/risk clearly defined, valid, reliable, and implemented consistently (including the same time period) across all study participants? 11. Were the assessors of exposure/risk blinded to the case or control status of participants? 12. Were key potential confounding variables measured and adjusted statistically in the analyses? If matching was used, did the investigators account for matching during study analysis?

+, criteria satisfied; - , not satisfied; N/A, not applicable; CD, cannot determine; * non peer-reviewed preprint study

| Author, year | Study design | N Treatment/ Control | Outcome recorded (day) | Control | | | | Intervention | | | |
|---|---|---|---|---|---|---|---|---|---|---|---|
| | | | | Dead | Ventilated | Hospitalised | Discharged | Dead | Ventilated | Hospitalised | Discharged |
| **ANAKINRA** | | | | | | | | | | | |
| Balkhair, 2020 | Prospective with control | 45/24 | 14 | 2 | 11 | 5 | 6 | 5 | 9 | 6 | 25 |
| Huet, 2020 | Prospective with control | 52/44 | - | 32 # | - | - | - | 13 # | - | - | - |
| Kooistra, 2020 | Prospective with control | 21/39 | 28 | 7 | - | - | - | 4 | - | - | - |
| *Kyriazopoulou, 2020 | Prospective with control | 130/130 | 30 | 16 | - | - | - | 6 | - | - | - |
| Cauchois, 2020 | Retrospective | 12/10 | 15 | 1 | 1 | 6 | 2 | 0 | 0 | 3 | 9 |
| Cavalli, 2020 | Retrospective | 29/16 | 21 | 7 | 1 | 1 | 7 | 3 | 5 | 8 | 13 |
| Narain, 2020 | Retrospective | 57/3076 | - | - | - | - | - | - | - | - | - |
| **SARILUMAB** | | | | | | | | | | | |
| Benucci, 2020 | Prospective | 8/0 | 14 | - | - | - | - | 1 | 0 | 0 | 7 |
| Della-Torre, 2020 | Prospective with control | 28/28 | 28 | 5 | 2 | 4 | 17 | 2 | 4 | 5 | 17 |
| * Gordon, 2021 | Adaptive RCT | 45/397 | 14 | Adjusted OR for improvement – 1.86 (95%CI 1.22-2.91) | | | | | | | |
| Gremese, 2020 | Prospective | 53/0 | 15 | - | - | - | - | 2 | 7 | 25 | 19 |
| Sinha, 2020 | Prospective | 255/0 | 25 | - | - | - | - | 28 | 1 | 9 | 218 |
| **SILTUXIMAB** | | | | | | | | | | | |
| * Gritti, 2020 | Prospective with cohort | 30/30 | 15 | - | - | - | - | 6 | 11 | 8 | 5 |
| **TOCILIZUMAB** | | | | | | | | | | | |
| Albertini, 2020 | Prospective with control | 22/22 | 14 | 0 | 6 | 14 | 2 | 1 | 4 | 16 | 1 |
| Antony, 2020 | Prospective | 80/0 | N/R | - | - | - | - | 7 | 9 | - | - |
| Campins, 2020 | Prospective | 58/0 | N/R | - | - | - | - | 8 | - | - | - |

| | | | | | | | | | | | |
|---|---|---|---|---|---|---|---|---|---|---|---|
| * Carvalho, 2020 | Prospective with control | 29/24 | 14 | 4 | - | - | - | 5 | - | - | - |
| Dastan, 2020 | Prospective | 42/0 | 15 | - | - | - | - | 6 | 6 | 11 | 19 |
| * Gordon, 2021 | Adaptive RCT | 350/397 | 14 | Adjusted OR for improvement – 1.83 (95%CI 1.40-2.41) | | | | | | | |
| Hermine, 2020 | Open label RCT | 63/67 | 14 | 6 | 11 | 20 | 30 | 7 | 3 | 21 | 32 |
| Malekzadeh, 2020 | Prospective | 126/0 | 14 | - | - | - | - | 24 | 9 | 7 | 86 |
| Mikulska, 2020 | Prospective with control | 29/66 | 14 | 16 | - | - | - | 4 | 2 | - | - |
| Morena, 2020 | Prospective | 51/0 | 15 | - | - | - | - | 14 | 2 | 35 | 0 |
| Perrone, 2020 | open-label phase 2 trial | 180/121 | 14 | 27 | - | - | - | 27 | - | - | - |
| Perrone, 2020 | Prospective with control | 528/360 | 14 | 45 | - | - | - | 56 | - | - | - |
| * Rosas, I., 2020 | phase 3 RCT | 294/144 | 28 | 28 | 23 | 22 | 71 | 50 | 44 | 26 | 166 |
| Roumier, 2020 | Prospective with control | 49/47 | 28 | 5 | - | - | 33 | 6 | - | - | 23 |
| Salama, 2020 | Double-blind RCT | 249/128 | 28 | 11 | - | - | - | 26 | - | - | - |
| Salvarani, 2020 | Open label RCT | 60/63 | 14 | 1 | 5 | 21 | 36 | 1 | 6 | 19 | 34 |
| * Sanchez-Montalva, 2020 | Prospective | 82/0 | 7 | - | - | - | - | 22 | 14 | 12 | 34 |
| Sciascia, 2020 | Prospective | 63/0 | 14 | - | - | - | - | 7 | 2 | - | - |
| Stone, 2020 | Double blind RCT | 161/82 | 28 | 3 | - | - | 72 | 9 | - | - | 147 |
| Strohbehn, 2020 | Phase 2 open label | 32/41 | 28 | - | - | - | - | 5 | - | - | - |
| Toniati, 2020 | Prospective | 100/0 | 10 | - | - | - | - | 20 | - | - | 15 |
| Biran, 2020 | Retrospective | 210/420 | N/R | - | - | - | - | 102 | - | - | 135 |
| Canziani, 2020 | Retrospective | 64/64 | N/R | 24 | - | - | - | 17 | - | - | - |
| Capra, 2020 | Retrospective | 62/23 | 9 | 11 | 4 | 0 | 8 | 2 | 5 | 32 | 23 |
| Chillmuri, 2020 | Retrospective | 83/685 | N/R | - | - | - | - | - | - | - | - |
| De Rossi, 2020 | Retrospective | 90/68 | N/R | 34 | 6 | - | - | 7 | 13 | - | - |
| Eimer, 2020 | Retrospective | 22/22 | 30 | 7 | 5 | 7 | 3 | 5 | 1 | 4 | 12 |

| | | | | | | | | | | | |
|---|---|---|---|---|---|---|---|---|---|---|---|
| Fisher, 2020 | Retrospective | 45/70 | 30 | 28 | - | - | - | 13 | - | - | - |
| Galvan Roman, 2020 | Retrospective | 58/88 | 61 | 16 | - | - | - | 14 | - | - | - |
| * Garcia, 2020 | Retrospective | 77/94 | 14 | - | - | - | 71 | - | - | - | 65 |
| Gokhale, 2020 | Retrospective | 70/91 | N/R | 61 | - | - | 30 | 33 | 2 | 9 | 26 |
| Guaraldi, 2020 | Retrospective | 179/365 | 14 | 60 | 117 | - | - | 9 | 42 | - | - |
| Guisado-Vasco, 2020 | Retrospective | 132/475 | N/R | 97 | - | - | - | 44 | - | - | - |
| Gupta, 2020 | Retrospective | 433/3492 | 27 | 1419 | - | - | - | 125 | - | - | - |
| Hill, 2020 | Retrospective | 43/45 | 28 | 15 | 0 | 3 | 27 | 9 | 6 | 2 | 26 |
| Holt, 2020 | Retrospective | 24/30 | N/R | - | - | - | - | - | - | - | - |
| Ip, 2020 | Retrospective | 134/413 | 30 | 231 | - | - | - | 62 | - | - | - |
| Kewan, 2020 | Retrospective | 28/23 | 14 | 2 | 7 | 4 | 10 | 3 | 10 | 5 | 10 |
| Kimmig, 2020 | Retrospective | 54/57 | N/R | 11 | - | - | 34 | 19 | - | - | 18 |
| Klopfenstein, 2020 | Retrospective | 20/25 | N/R | 12 | - | - | 11 | 5 | - | - | 11 |
| Lewis, 2020 | Retrospective | 497/497 | N/R | 211 | - | - | 283 | 145 | - | - | 332 |
| Martinez-Sanz, 2020 | Retrospective | 260/969 | N/R | 120 | - | - | - | 61 | - | - | - |
| Narain, 2020 | Retrospective | 73/3076 | N/R | - | - | - | - | - | - | - | - |
| Nasa, 2020 | Retrospective | 22/63 | 28 | 36 | - | - | - | 2 | - | - | - |
| Patel, 2020 | Retrospective | 42/41 | 7 | 11 | - | 7 | 7 | 9 | - | | 7 |
| * Petrak, 2020 | Retrospective | 81/37 | N/R | - | - | - | - | - | - | - | - |
| Pettit, 2020 | Retrospective | 74/74 | N/R | 17 | - | - | - | 29 | - | - | - |
| Potere, 2020 | Retrospective | 10/10 | 14 | 0 | 1 | 4 | 5 | 0 | 0 | 2 | 8 |
| * Ramaswamy, 2020 | Retrospective | 21/65 | N/R | 8 | - | - | - | 3 | - | - | - |
| Rodriguez-Bano, 2020 | Retrospective | 88/344 | 21 | 41 | 20 | 30 | 253 | 2 | 6 | 10 | 70 |
| Rojas-Marte, 2020 | Retrospective | 96/97 | N/R | 55 | - | - | - | 43 | - | - | |
| Roomi, 2020 | Retrospective | 32/144 | N/R | 13 | - | - | 38 | 6 | - | - | 25 |
| Rosas,J., 2020 | Retrospective | 20/17 | 30 | 6 | - | - | - | 4 | - | - | - |
| Rossi, 2020 | Retrospective | 84/84 | N/R | - | - | - | - | - | - | - | - |
| Rossotti, 2020 | Retrospective | 74/148 | NR | - | - | - | - | 8 | 18 | 45 | 14 |
| Ruiz-Antoran, 2020 | Retrospective | 268/238 | N/R | 75 | - | - | - | 45 | - | - | - |
| Somers, 2020 | Retrospective | 78/76 | 14 | 28 | 15 | 11 | 22 | 14 | 21 | 12 | 31 |
| Tian, 2020 | Retrospective | 65/130 | N/R | 42 | - | - | - | 14 | - | - | - |
| Tsai, 2020 | Retrospective | 66/66 | N/R | 18 | - | - | - | 18 | - | - | - |

| | | | | | | | | | | | |
|---|---|---|---|---|---|---|---|---|---|---|---|
| * Wadud, 2020 | Retrospective | 44/50 | N/R | 26 | - | - | - | 17 | - | - | - |
| Zheng, 2020 | Retrospective | 92/89 | 27.5 | 1 | 0 | 0 | 88 | 9 | 0 | 0 | 83 |

**Supplementary Table 4** – Primary clinical outcome. Outcome scores presenting using absolute scores with number of individuals in each category, using adapted ordinal outcome scores 1 indicates death, 2 described hospitalised patients requiring invasive ventilatory support, 3 describes patients not requiring invasive ventilatory support but still hospitalised, 4 describes discharged patients. Day outcomes reported shown where applicable.

* non peer-reviewed preprint study, CI, confidence interval

### death or ventilation

| Retrospective studies | | | | | | | | |
|---|---|---|---|---|---|---|---|---|
| Variables | Generalised odds ratios for ordinal outcomes (N=10) | | Difference in duration of hospitalisation (N=9) | | Adjusted hazard ratios for mortality (N=18) | | Risk ratios for mortality (N=31) | |
| | $R^2$ | P value | $R^2$ | P value | $R^2$ | P value | $R^2$ | P value |
| Steroid use | 0.00 | 0.7921 | 7.17 | 0.2305 | 0.00 | 0.9710 | 0.00 | 0.5252 |
| Peer review | N/A | N/A | N/A | N/A | 0.9 | 0.5638 | 0.00 | 0.4137 |
| Route of administration | 4.75 | 0.3526 | 81.64 | <0.001 | 0.00 | 0.3921 | 2.68 | 0.2053 |
| Single centre | 0.00 | 0.6028 | 11.03 | 0.2013 | 2.96 | 0.4103 | 0.00 | 0.2154 |
| Outcome day | 0.00 | 0.7921 | N/A | N/A | 99.99 | 0.0818 | 9.54 | 0.4141 |
| Prospective studies | | | | | | | | |
| Variables | Generalised odds ratios for ordinal outcomes (N=5) | | Difference in duration of hospitalisation (N=1) | | Adjusted hazard ratios for mortality (N=4) | | Risk ratios for mortality (N=11) | |
| | $R^2$ | P value | $R^2$ | P value | $R^2$ | P value | $R^2$ | P value |
| Steroid use | 99.99 | <0.0001 | N/A | N/A | 45.29 | 0.3464 | 0.00 | 0.9050 |
| Peer review | 0.00 | 0.4890 | N/A | N/A | N/A | N/A | 0.00 | 0.5764 |
| Route of administration | N/A | N/A | N/A | N/A | 45.29 | 0.3464 | 69.89 | 0.5922 |
| Single centre | 0.00 | 0.5332 | N/A | N/A | 0.00 | 0.2425 | 0.00 | 0.8638 |
| Outcome day | 0.00 | 0.5351 | N/A | N/A | 0.00 | 0.7187 | 0.00 | 0.6115 |

**Supplementary Table 5 -** Results of meta-regression for variables assessed separated by study design (retrospective and prospective) and study outcomes. Study numbers for each outcome shown (N). $R^2$ and p values from meta-regression shown were applicable.
N/A, not applicable.

| Outcome | The GRADE domains | Ratings for quality of evidence |
|---|---|---|
| Ordinal scale (12 studies; 4 prospective and 8 retrospective. Total of 1782 patients) | Risk of bias | Of the 4 prospective included, 3 RCTs of low/moderate risk of bias included. Retrospective studies generally of fair quality, although cannot exclude failure to control confounding factors. |
| | Imprecision | No serious imprecision, with appropriately narrow 95% confidence intervals. Outcome based on 1782 patients. |
| | Inconsistency | High inconsistency with significant heterogeneity in both prospective and retrospective studies. |
| | Indirectness | No serious indirectness. All studies included a control arm from the same population. All study subjects had Covid-19, although severity and participation criteria were inconsistent. |
| | Publication bias | No publication bias as indicated by funnel plots and Egger's tests |
| | Certainty of evidence | Moderate certainty of evidence. |
| Difference in duration of hospitalisation (9 retrospective studies, 1 RCT. Total of 2285 patients) | Risk of bias | All included retrospective studies with moderate/high risk of bias. Confounding factors were poorly controlled for. |
| | Imprecision | Serious imprecision, with studies showing shorter and longer duration of hospitalisation with tocilizumab. Appropriately narrow 95% confidence intervals. |
| | Inconsistency | High inconsistency with significant heterogeneity ($I^2$ = 93.8%). |
| | Indirectness | No serious indirectness. All studies included a control arm from the same population. All study subjects had Covid-19, although severity and participation criteria were inconsistent. |
| | Publication bias | No publication bias as indicated by funnel plots and Egger's tests |

| | | |
|---|---|---|
| | Certainty of evidence | Low certainty of evidence. |
| Overall mortality (aHR - 22 studies. Total of 13,702 patients. RR - 42 studies, 15,085 patients) | Risk of bias<br>Imprecision<br>Inconsistency<br>Indirectness<br>Publication bias<br>Certainty of evidence | RCTs of low/moderate risk of bias included.<br>No imprecision present<br>High inconsistency in retrospective studies, but not in prospective studies.<br>No serious indirectness. All studies included a control arm from the same population. All study subjects had Covid-19, although severity and participation criteria were inconsistent<br>No publication bias as indicated by funnel plots and Egger's tests<br>High certainty of evidence. |

**Supplementary Table 6 –** GRADE (Grading of Recommendations, Assessment, Development and Evaluations) approach to rate the quality of evidence on the effects of tocilizumab
